# Mesial-to-lateral patterns of epileptiform activity identify the seizure onset zone in mesial temporal lobe epilepsy

**DOI:** 10.1101/2024.10.29.24316309

**Authors:** Carlos A. Aguila, Alfredo Lucas, Sarah Lavelle, Akash R. Pattnaik, Juri Kim, William K.S. Ojemann, Devin Ma, Mariam Josyula, Nina Petillo, Joshua J. LaRocque, Saurabh R. Sinha, Colin A. Ellis, Alexandra Parashos, Ezequiel Gleichgerrcht, Kathryn A. Davis, Brian Litt, Erin C. Conrad

**Affiliations:** Department of Bioengineering, School of Engineering & Applied Sciences, University of Pennsylvania, Philadelphia, PA 19104, USA; Center for Neuroengineering and Therapeutics, University of Pennsylvania, Philadelphia, PA 19104, USA; Department of Neurology, Perelman School of Medicine, University of Pennsylvania, Philadelphia, PA 19104, USA; Perelman School of Medicine, University of Pennsylvania, Philadelphia, PA 19104, USA; Department of Neurology, Emory University, Atlanta, GA 30325, USA; Department of Neurology, Medical University of South Carolina, Charleston, SC 29425 USA

## Abstract

Mesial temporal lobe epilepsy (mTLE) is a common localization of drug-resistant epilepsy in adults. Patients often undergo intracranial EEG (iEEG) monitoring to confirm localization and determine candidacy for focal ablation or resection. Clinicians primarily base surgical decision-making on seizure onset patterns, with imaging abnormalities and information from interictal epileptiform discharge (spikes) used as ancillary data. How the morphology and timing of spikes within multi-electrode sequences may inform surgical planning is unknown, in part due to the lack of measurement methods for large datasets. We hypothesized that patients with mTLE have a distinct mesial-to-lateral spike pattern that differentiates them from other epilepsy localizations. In a multicenter study at the University of Pennsylvania and the Medical University of South Carolina, we analyzed the timing and morphology of spikes and seizure high frequency energy ratio (HFER) in 75 patients with drug-resistant epilepsy. We compared these features across patients with mTLE, temporal neocortical epilepsy, and other localizations. A logistic regression model combining all features predicted a clinical localization of mTLE in unseen patients with an AUC of 0.82 (compared to an AUC of 0.70 for seizure-only features, DeLong’s test *p* = 0.08). Spike rate was the most important feature in the combined model. Modeled probability of mTLE was similar between patients who had a good versus a poor 12-month outcome after undergoing a mesial temporal resection or ablation (*p* = 0.34). These findings support quantitative spike analysis to supplement analysis of seizures for use in surgical planning.

## Introduction

The mesial temporal lobe is a common site of seizure onset in patients with drug-resistant epilepsy. Distinguishing mTLE from other epilepsy localizations is crucial, as it can be treated with specific minimally-invasive surgical procedures such as laser ablation. Less invasive procedures are often preferred over a standard temporal lobectomy due to higher surgical risks and increased chances of cognitive and focal neurological deficits in the latter procedure (Donos et al., 2018; Helmstaedter, 2008; Seidenberg et al., 1998; Spencer & Huh, 2008). However, the success rates of mesial temporal laser ablation are suboptimal, with ∼50% of patients achieving seizure freedom after 12 months (Gross et al., 2018), compared to ∼65% for anterior temporal lobectomies (Engel et al., 2012; Wiebe et al., 2001). This disparity in efficacy highlights the need for improved methods to identify patients who have mTLE, and thus who would be good candidates for targeted ablations.

Current surgical planning relies on capturing seizures on intracranial EEG recordings and then visually identifying the seizure onset zone (SOZ). However, this process is error-prone, hindered by low interrater reliability (Davis et al., 2018) and challenges in precise electrode positioning (Conrad, Bernabei, et al., 2020). Furthermore, prolonged hospital stays with multiple seizures carry a risk of morbidity (Hamer et al., 2002; Johnson, 2019; Sperling, 1997). There is a pressing need for better methods to localize mTLE that can both shorten hospital stays and improve surgical outcomes.

Interictal spikes — brief electrical discharges observed between seizures — may also localize the SOZ. Spikes are thought to originate from seizure-generating regions, as supported by human studies in source localization using spikes (Mitsuhashi et al., 2021; Withers et al., 2023) and animal model studies (Bink et al., 2018; Vakharia et al., 2018). To supplement seizure data, epileptologists often visually analyze spikes, looking both at spike frequency and the timing of spikes in multi-channel sequences as potential markers of the SOZ (Bautista et al., 1999; Rodin et al., 2009). However, spikes play a relatively minor role in surgical planning. This is in part because of the evidence that many spikes occur outside the SOZ (Hamer et al., 1999; Lüders et al., 2006; Ramantani et al., 2023), but also because our current approach of visual spike analysis is only feasible when performed on a small number of spikes. New advances in spike detection and feature extraction algorithms now allow us to parse the large datasets acquired in intracranial EEG monitoring, and to study the precise timing and morphology of spikes in multi-channel spike sequences. Prior studies have found that earlier spikes in sequences (Hufnagel et al., 2000; Khoo et al., 2018) and those with sharper, larger shapes (Serafini, 2019; Thomas et al., 2023) are closer to the epileptogenic source. It remains unclear how combining features of spike rate, timing, and morphology adds to our ability to localize the epileptogenic cortex, or what this information adds beyond seizure data.

We hypothesize that in mTLE, spikes originate in the mesial temporal lobe, and thus have a consistent mesial-to-lateral pattern when observed on electrodes targeting the mesial temporal structures in a mesial-to-lateral trajectory(Gompel et al., 2010) **(Fig 1A)**. We predict mesial-to-lateral patterns in spike rates, timing in multi-contact sequences, and morphology that distinguish mTLE from other epilepsy localizations. To test this hypothesis, we studied patients with drug-resistant epilepsy who underwent intracranial EEG monitoring. We measured the mesial-to-lateral patterns of spike features and seizure high-frequency activity in temporal lobe electrodes. This quantitative analysis of spike rate, morphology, and timing can be used to supplement seizure data to localize the epileptogenic zone, potentially reducing the number of seizures required to make accurate diagnoses. This approach aims to enhance our understanding of mTLE and improve surgical planning and outcomes.

**Figure 1.**
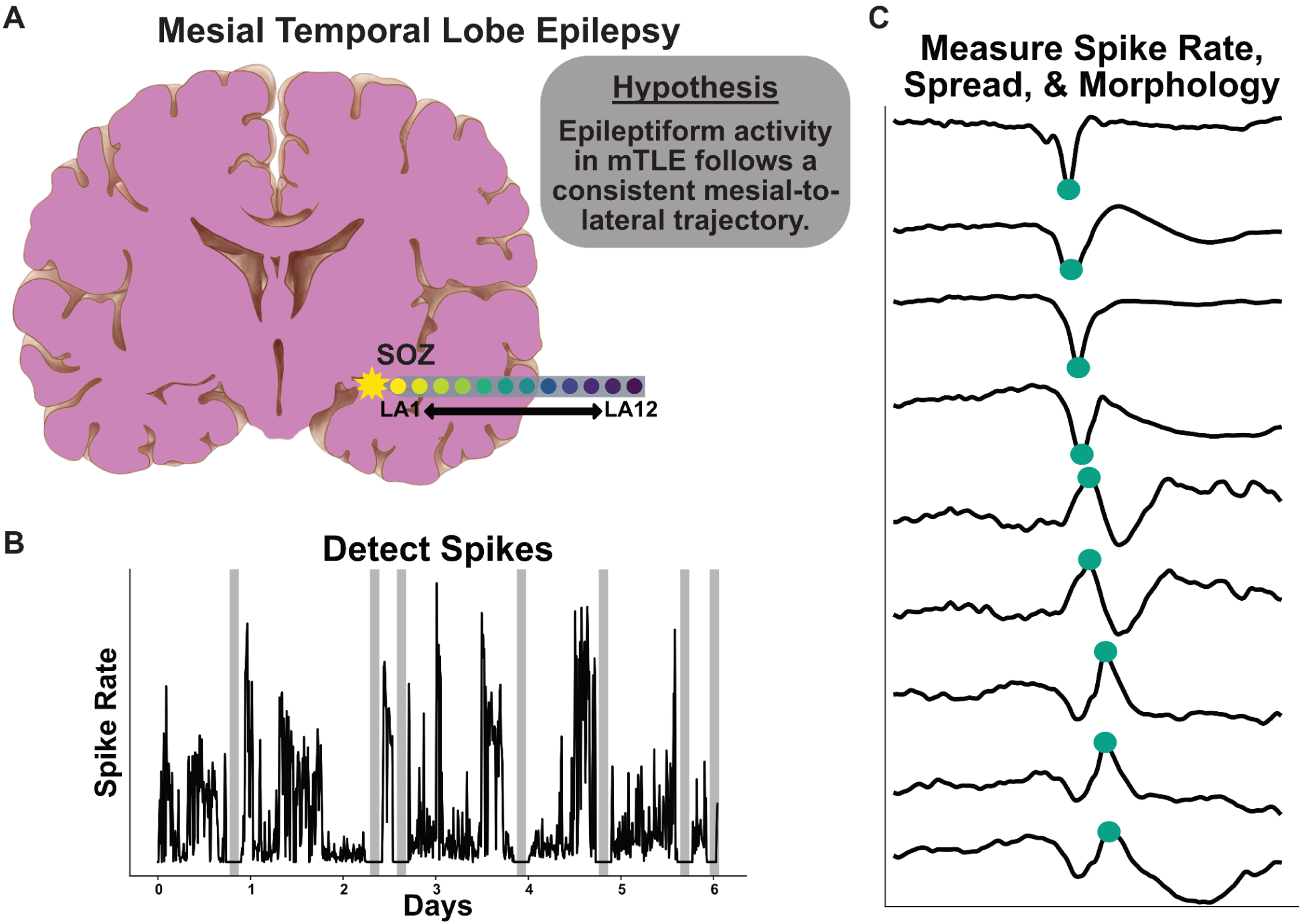
Hypothesis and methodology for investigating mesial-to-lateral epileptiform activity in mesial temporal lobe epilepsy (mTLE). (A) Schematic representation of the hypothesized mesial-to-lateral trajectory of epileptiform activity in mTLE. The electrode array (colored circles) spans from the SOZ to lateral temporal regions (LA1 to LA12). (B) Representative intracranial EEG recording demonstrating spike detection over multiple days, with varying spike rates indicated by amplitude fluctuations. Gray bars are periods of seizure activity. (C) Example of spike train detection across multiple channels. This demonstrates our ability to identify spike trains, from which we can extract timing, rate, and morphological features (green markers). The progression from top to bottom may represent different electrode contacts, allowing for analysis of spatial and temporal patterns in epileptiform activity.

## Patients and Methods

### Patient Selection and Clinical Determinations

Research data collection had prior approval from the institutional review boards at the Hospital of the University of Pennsylvania (HUP) and the Medical University of South Carolina (MUSC). All participants provided informed consent. We included patients with drug-resistant focal epilepsy who underwent intracranial EEG (iEEG) recording in the Epilepsy Monitoring Unit (EMU) as part of surgical planning (HUP date range: 2013-2021; MUSC date range 2021-2022), consented to analysis of their intracranial data, and who had depth electrode coverage of temporal lobe structures ipsilateral to the clinician-defined seizure onset zone. Seizure times and the anatomical seizure onset localization were identified by a board-certified epileptologist as part of clinical care, and confirmed in a clinical case conference consisting of multiple epileptologists. We designated epilepsy localizations as mesial temporal, temporal neocortical, or “other” cortex. We excluded patients who did not have a clear epileptic focus (multifocal or diffuse epilepsy). For patients who underwent resection or ablation specifically targeting the mesial temporal structures, one-year Engel scores were determined by the board-certified neurologist treating the patient (Engel, 1993; Epilepsy (ILAE) 1997–2001 et al., 2001).

### Interictal iEEG data selection and pre-processing

We examined all interictal iEEG data, excluding time periods within two hours of a seizure. The continuous interictal periods were divided into 10-minute intervals, and within each interval, a one-minute epoch was randomly selected. Automatic processing of interictal spikes for each epoch was accomplished using a previously validated, in-house spike detector (Brown et al., 2007; Conrad, Tomlinson, et al., 2020). A board-certified epileptologist (EC) visually analyzed a random sample of 50 spike detections for each patient and determined how many appeared to be real spikes as opposed to artifact or non-spike cerebral activity. We excluded patients for which the positive predictive value (PPV) of spike detection accuracy was less than 75%. To ensure adequate spatial sampling, only electrodes with at least 8 contacts in the brain were included in the analysis.

### Feature Calculation

#### Interictal

We computed a set of morphological, rate, and timing features for each spike. Standard morphology characteristics were calculated for both the sharp, initial wave portion and the subsequent slow wave (Y.-C. Liu et al., 2013; Thomas et al., 2023). Detailed calculations for each feature can be found in the Supplemental Materials. First, we measured the spike rate for each electrode contact, defined as the average number of spikes divided by the total duration **(Fig 1B)**. Morphological features included sharp wave rising and falling amplitude, sharp wave rising and falling slope, sharpness (sum of the absolute value of slopes), rising and falling width, line length, slow wave amplitude, and slow wave width. We also measured the *timing* of electrode contacts involved in *spike trains —* sequences of spikes occurring in close succession but with discernable latency —, representing the latency between the start of a spike train and the time a spike is detected on each electrode contact **(Fig 1C)**. Morphological features were extracted for spikes detected in each epoch and concatenated across the entire interictal interval. Spike rate, timing, and morphology were averaged across all spikes within an individual electrode contact. The Supplemental Materials describes our approach to examining the consistency of spike sequence order over time.

#### Seizure

We analyzed iEEG seizure data from patients, with ictal start and stop times annotated by a board-certified epileptologist. Raw signals were filtered into high-gamma frequency bands (70-140Hz) using a fourth-order Butterworth IIR filter, with a 60Hz notch filter to remove power line noise (Cai et al., 2022; Virtanen et al., 2020). A 60s baseline segment was extracted for each seizure, ending 40s before ictal onset (Cai et al., 2022). We computed seizure high frequency energy ratios (HFER) for all seizures. This metric was adapted from the epileptogenicity index (EI), an index for measuring the fast oscillation epileptogenicity on each channel, by omitting the channel onset scaling portion of EI due to poor accuracy in automated seizure start detection (Bartolomei et al., 2008; Cai et al., 2022). The Supplemental Materials fully describes our approach to measuring seizure HFER. Briefly, we identified clinician-defined seizure onset times and identified target periods to be 1 seconds before and 10 seconds after seizure onset, chosen to maximize likelihood of capturing meaningful seizure spread associated with good seizure outcomes (Andrews et al., 2020; Revell et al., 2022). We tested multiple target window lengths (6s, 11s, 16s, and 21s) to measure the sensitivity of ictal features to this result. The HFER was calculated as the ratio of high-frequency (70-140Hz) energy in the target window to that in a baseline window (100-40s pre-seizure). HFER values were normalized using the mean and standard deviation across all channels for each seizure. We then took the median normalized HFER across all seizures, resulting in a single HFER for each electrode contact. Intuitively, an electrode with a higher HFER has disproportionately more high frequency energy relative to the interictal baseline than other electrodes at the start of seizures.

### Measuring the spatial distribution of spike and seizure features along temporal lobe electrode contacts

Given our hypothesis that spikes in mTLE originate in the mesial temporal structures, we predicted that patients with mTLE would have disproportionately more frequent, high amplitude, sharper spikes, with earlier timing in multi-spike sequences in the mesial temporal structures compared to patients with other epilepsy localizations. To test this, we restricted analysis to electrodes targeting the temporal lobe structures in a mesial-to-lateral fashion. We chose to study this particular electrode trajectory because this is a common approach used to sample the mesial temporal structures at both HUP and MUSC. To approximate the mesial-to-lateral spatial distribution of each feature, we measured the *mesial-to-lateral spread*, which we defined as the Pearson correlation between the feature across these electrodes with a vector ranging from 1 to the number of contacts on the electrode (which varied in number between 8-12). If patients had bilateral epilepsy, features were averaged across same-numbered contacts on both sides. This vector represents the mesial-to-lateral axis. Our prediction of a greater mesial-to-lateral pattern of spikes would yield a prediction of *positive* Pearson correlations for some analyses, and *negative* correlations for others. For example, we would expect a *negative* Pearson correlation for spike rates, implying that higher-numbered, or more lateral, contacts have lower spike rates; but a *positive* correlation for spike timing, implying that higher-numbered contacts have later timing of spikes in multi-channel sequences. We performed the same analysis to calculate the mesial-to-lateral spread of seizure HFER. We compared the mesial-to-lateral spread for each feature between patients with mesial temporal, temporal neocortical, and other epilepsy localizations.

### Machine Learning to predict mTLE

We next tested whether the mesial-to-lateral spread of spike and seizure HFER could predict a localization of mTLE in unseen patients. Features included spike rate, spike timing, rising and falling amplitude, rising and falling slope, sharpness, line length, spike width, slow wave amplitude, slow wave width, and seizure HFER. We performed classification using Elastic Net Logistic Regression to predict a localization of mTLE vs. either temporal neocortical epilepsy or other cortical localizations (we combined the latter two localizations into a single class to have a binary classification problem for identifying mTLE). Our classifier was trained, tested, and tuned inside a 2-loop model. The outer fold consisted of a Leave One Out Cross Validation (LOOCV) model, where we trained our classifier on all patients except for one, which was left out for testing. This process was repeated until all patients were tested. The inner fold consisted of a 5-fold cross validation grid search algorithm that maximized our classifying accuracy by tuning the number of iterations to solve the gradient, the alpha value, and the ratio of L1 to L2 penalty. To understand how spike morphology and timing features provided additional localizing value over seizures, we also constructed a model trained solely on the mesial-to-lateral pattern of seizure HFER. Feature importance was computed by taking the average of the coefficients of the logistic regression model across all classes and training folds. Balanced accuracy was also calculated by taking the average of recall obtained on each class.

### Associating spike feature spatial distribution with surgical outcome after mesial temporal ablation

We hypothesized that patients with mTLE would have both 1) a stronger mesial-to-lateral pattern of spike and seizure features, and 2) better surgical outcomes after undergoing surgery targeting the mesial temporal lobe structures. To test this, we compared the predicted probabilities of mTLE from our machine learning model between patients who had a good (Engel Class I) versus a poor (Engel 2+) 12-month outcome after undergoing mesial temporal laser ablation (Jaber et al., 2024). We also created logistic regression models to predict patient outcomes directly from their spike, seizure, or combined features.

### Statistical analyses

To compare two independent groups, we report a Mann-Whitney U test (alpha = 0.05) and Cohen’s d for effect sizes, where values of 0.20, 0.50 and 0.80 were considered as small, medium and large effects, respectively. To compare more than two groups, we report Kruskil-Wallis tests. Classification results were presented using receiver operating characteristics curve (ROC), area under ROC (AUC), and balanced accuracy. We used DeLong’s test to compare AUCs across our different models. Analyses were performed in Python 3.12.0. For all assessments, the significance threshold (α) was set at 0.05 (Benjamini-Hochberg corrected for multiple comparisons), and significance is indicated in figures with * = p < 0.05, ** = p < 0.01, *** = p < 0.001, and **** = p < 0.0001.

### Data and code availability

Raw EEG data is available on ieeg.org. All code used to perform analyses, along with an intermediate dataset containing electrode contact-level features, is publicly available on https://github.com/penn-cnt/IEEG_Spike-Morphology. A readme document outlines the necessary steps to initialize the analyses. Complete patient iEEG data is available on iEEG.org where users can access free of charge. All patients are part of the *HUP_Intracranial_Data* project on iEEG.org and available on request.

## Results

### Patient information

75 patients were included in our analysis (51 from HUP, 24 from MUSC). 14 patients, all from HUP, were excluded due to low accuracy spike detections. **Table 1** contains a summary of the patient demographics for this study. 49 were diagnosed with mTLE, 8 with temporal neocortical epilepsy, and 18 with epilepsy arising from other cortical regions. A median (IQR) of 24632 (10723 - 60247) spikes were detected across patients. The median (IQR) of spike detector accuracy as measured by the PPV was 0.90 (0.84-0.96).

**Table 1.**
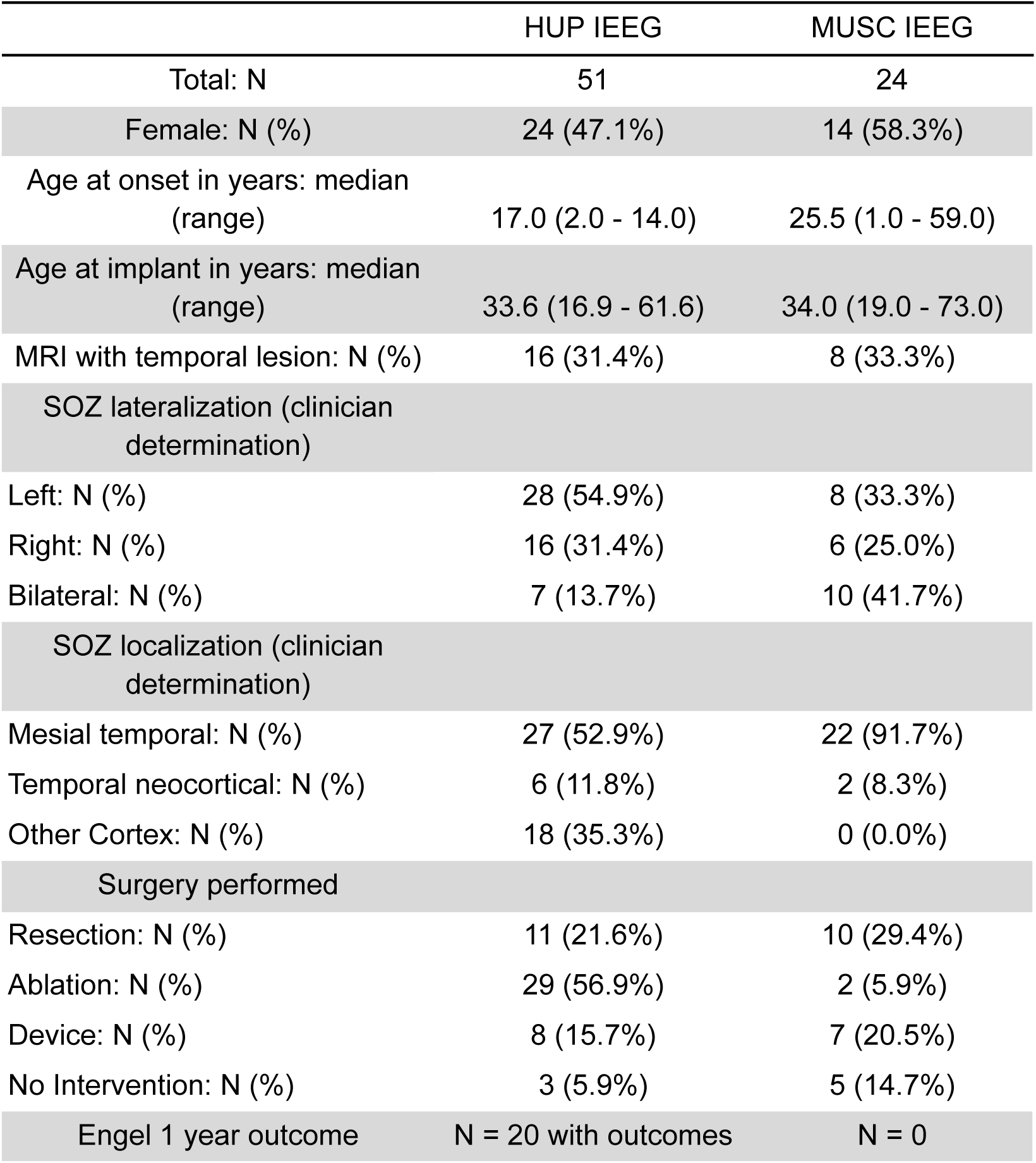
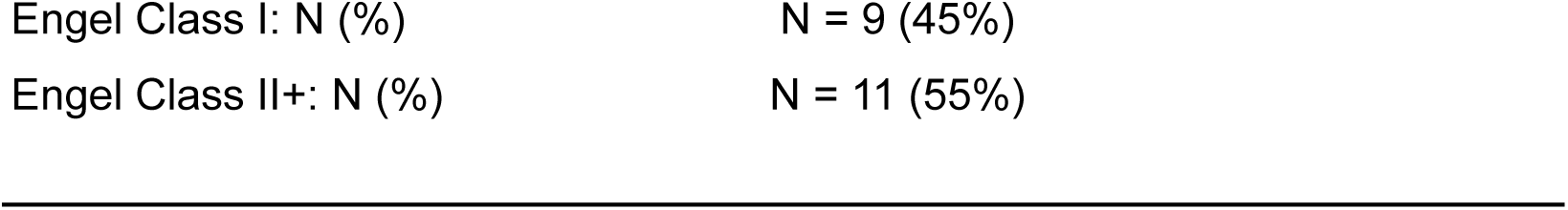
Demographic and clinical summary statistics on patients from HUP and MUSC cohorts. 12-month Engel outcomes are for patients who underwent a mesial temporal lobe ablation.

|  | HUP IEEG | MUSC IEEG |
| --- | --- | --- |
| Total: N | 51 | 24 |
| Female: N (%) | 24 (47.1%) | 14 (58.3%) |
| Age at onset in years: median (range) | 17.0 (2.0 - 14.0) | 25.5 (1.0 - 59.0) |
| Age at implant in years: median (range) | 33.6 (16.9 - 61.6) | 34.0 (19.0 - 73.0) |
| MRI with temporal lesion: N (%) | 16 (31.4%) | 8 (33.3%) |
| SOZ lateralization (clinician determination) |  |  |
| Left: N (%) | 28 (54.9%) | 8 (33.3%) |
| Right: N (%) | 16 (31.4%) | 6 (25.0%) |
| Bilateral: N (%) | 7 (13.7%) | 10 (41.7%) |
| SOZ localization (clinician determination) |  |  |
| Mesial temporal: N (%) | 27 (52.9%) | 22 (91.7%) |
| Temporal neocortical: N (%) | 6 (11.8%) | 2 (8.3%) |
| Other Cortex: N (%) | 18 (35.3%) | 0 (0.0%) |
| Surgery performed |  |  |
| Resection: N (%) | 11 (21.6%) | 10 (29.4%) |
| Ablation: N (%) | 29 (56.9%) | 2 (5.9%) |
| Device: N (%) | 8 (15.7%) | 7 (20.5%) |
| No Intervention: N (%) | 3 (5.9%) | 5 (14.7%) |
| Engel 1 year outcome | N = 20 with outcomes | N = 0 |

|  |  |
| --- | --- |
| Engel Class I: N (%) | N = 9 (45%) |
| Engel Class II+: N (%) | N = 11 (55%) |

### The mesial-to-lateral pattern of spike features and seizure HFER is distinct across epilepsy localizations

We predicted that patients with mTLE would have a mesial-to-lateral pattern of higher spike rates, earlier timing of spikes in multi-spike sequences, and higher-amplitude and sharper spikes in the mesial temporal structures; whereas, we expected this pattern to be either reversed or non-existent for patients with temporal neocortical or other localizations. We first examined spike rates, predicting that in mTLE, the mesial contacts would have higher spike rates, yielding a *negative* Pearson correlation with the mesial-to-lateral axis **(Fig 2A)**. We found a difference in mesial-to-lateral spread between epilepsy localizations (Kruskal-Wallis: χ²(2) = 27.64, p < 0.001). Post-hoc Mann-Whitney U tests revealed that patients with mTLE had significantly lower Pearson correlations (*r* = -0.88 (-0.82 - -0.92)) than patients with other-cortex localizations (*r* = -0.14 (-0.52 - 0.17); *U* = 72, Benjamini-Hochberg-corrected *p* < 0.001, Cohen’s *d* = 2.31), implying a stronger mesial-to-lateral pattern and agreeing with our prediction. Other pairs of localizations were not significantly different (B-H-corrected *p* > 0.05) **(Fig 2B)**.

**Figure 2.**
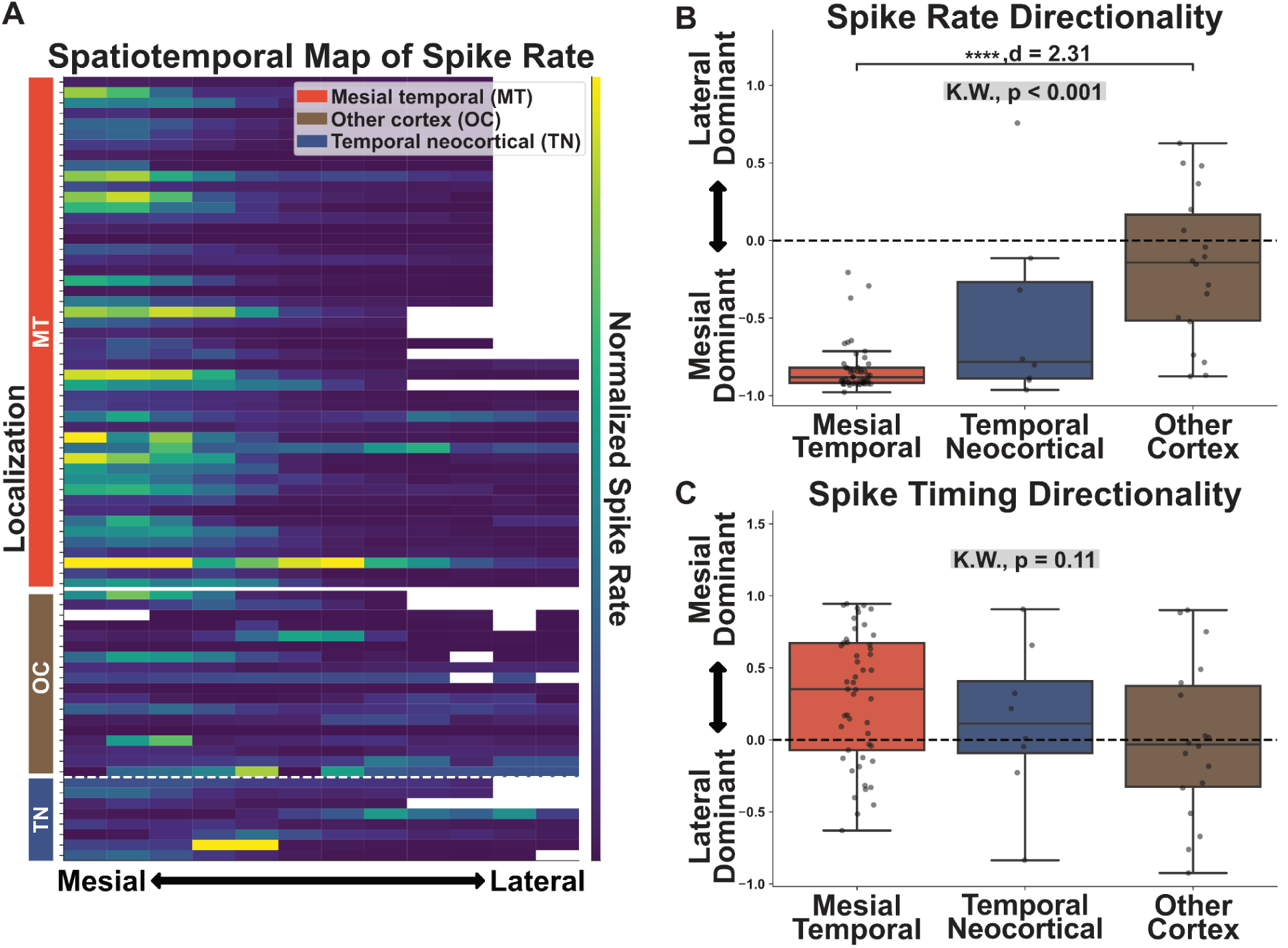
Mesial-to-lateral spread of spike rate and spike timing across epilepsy localizations. **(A)** Spatiotemporal heat map of spike rates. Each row represents an individual patient, with the colored bar on the left indicating epilepsy type: mesial temporal (MT, red), other cortex (OC, brown), or temporal neocortical (TN, blue). Columns represent electrode contacts arranged from mesial (left) to lateral (right), with color intensity denoting spike rate (purple: low, yellow: high). **(B)** Box plots showing spike rate directionality across different epilepsy localizations. The y-axis shows the Pearson correlation between spike rate and the mesial-to-lateral axis. Positive values indicate lateral dominance, negative values suggest mesial dominance. A significant difference is observed between groups (Kruskal-Wallis test, p < 0.001), with a large effect size (d = 2.31) between mesial temporal and other cortex. **(C)** Box plots illustrating spike timing directionality across epilepsy localizations. The y-axis shows the Pearson correlation between spike timing and the mesial-to-lateral axis. No significant difference is found between groups (Kruskal-Wallis test, p = 0.11). In B and C, each dot represents an individual patient, with significant differences between SOZ types indicated by asterisks (* p < 0.05, ** p < 0.01, *** p < 0.001, **** p < 0.0001). This figure reveals a distinct mesial-to-lateral gradient in spike rate but not in spike timing across different epilepsy localizations.

We next compared the mesial-to-lateral spread of the timing of spikes in multi-contact sequences. We predicted that the more mesial (or lower-numbered) contacts of mTLE patients would have earlier spikes in multi-contact sequences, leading to a *positive* Pearson correlation of spike timing. We observed no significant difference in the mesial-to-lateral spike timing (Kruskal-Wallis, χ²(2) = 4.42, p = 0.11. There was a non-significant trend toward higher Pearson correlations, implying more mesial-to-lateral spike timing, in mTLE (*r* = 0.35 (-0.07 - 0.67)) **(Fig 2C)**.

We next compared the mesial-to-lateral pattern of spike morphology between localizations, predicting higher-amplitude and sharper spikes in the mesial contacts for mTLE, leading to *negative* Pearson correlations of rising amplitude and spike sharpness. This subset of features was chosen based on the least correlated morphological features **(Supplemental Fig 4)**. Additional morphological features and their analyses are reported in the supplemental materials **(Fig 3A)**. We found a difference in mesial-to-lateral spread between epilepsy localizations for rising amplitude (Kruskal-Wallis: χ²(2) = 7.39, B-H corrected p = 0.04) and spike sharpness (Kruskal-Wallis: χ²(2) = 11.71, B-H corrected p < 0.01). Post-hoc Mann-Whitney U tests revealed that patients with mTLE had significantly lower Pearson correlations than patients with other-cortex localizations for rising amplitude (mTLE: *r* = -0.61 (-0.80 - -0.24), other-cortex: *r* = -0.32 (-0.48 - 0.22); B-H corrected *U* = 251, *p* < 0.01, Cohen’s *d* = 0.88) and spike sharpness (mTLE: *r* = -0.63 (-0.81 - -0.38), other-cortex: *r* = -0.08 (-0.53 - 0.14); B-H corrected *U* = 217, *p* < 0.01, Cohen’s *d* = 1.01), implying a stronger mesial-to-lateral gradient of spike morphology in mTLE patients, consistent with our hypothesis of preferential spike generation in mesial temporal structures in mTLE. Other pairs of localizations were not significantly different (B-H-corrected *p* > 0.05) **(Fig 3B)**.

**Figure 3.**
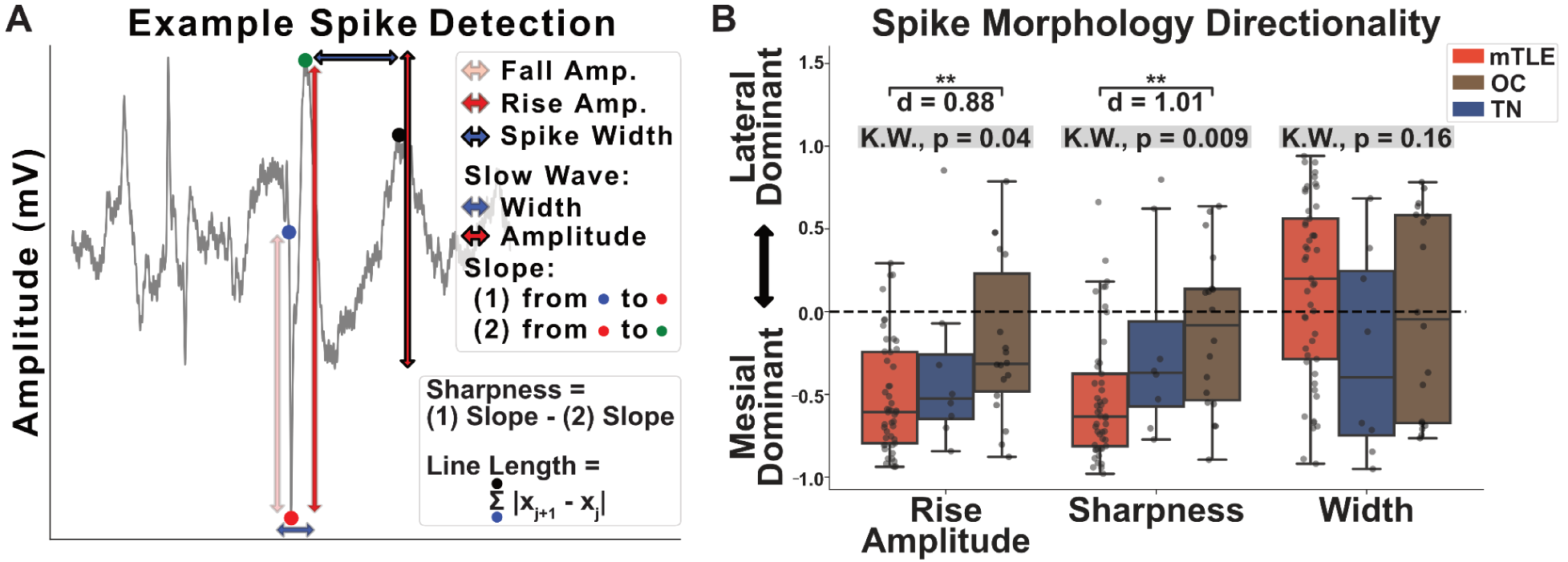
Mesial-to-lateral pattern of spike morphology across epilepsy localizations. **(A)** Example of spike detection and quantification of morphological features. The panel illustrates key parameters measured for each spike, including rise and fall amplitudes, spike width, and slow wave characteristics. Spike sharpness is defined as the difference between rise and fall slopes. **(B)** Box plots illustrate the directionality of spike morphology features across mTLE, other cortex (OC), and temporal neocortex (TN) epilepsy localizations. The y-axis shows the Pearson correlation between the morphological feature and the mesial-to-lateral axis. Rise amplitude, sharpness, and width are compared, with positive values indicating lateral dominance and negative values suggesting mesial dominance. Kruskal-Wallis (K.W.) tests reveal significant group differences for rise amplitude (p = 0.04) and sharpness (p = 0.009). Effect sizes (d) are provided for specific group comparisons: mTLE shows significantly different rise amplitude compared to OC (d = 0.88) and different sharpness compared to OC (d = 1.01). mTLE exhibits a distinct mesial-to-lateral gradient, particularly for rise amplitude and sharpness. Each dot represents an individual patient, with significant differences between SOZ types indicated by asterisks (* p < 0.05, ** p < 0.01, *** p < 0.001).

Finally, the mesial-to-lateral spread of seizure HFER was compared between localizations. We predicted higher energy in the mesial contacts for mTLE, signifying *negative* Pearson correlations of HFER **(Fig 4A)**. We found a difference in mesial-to-lateral spread between epilepsy localizations (Kruskal-Wallis: χ²(2) = 10.49, p < 0.01). Post-hoc Mann-Whitney U tests revealed that patients with mTLE had significantly lower Pearson correlations (*r* = -0.31 (-0.77 - 0.07) than patients with other-cortex localizations (*r* = 0.51 (0.02 - 0.69); *U* = 119, Benjamini-Hochberg-corrected *p* < 0.001, Cohen’s *d* = -1.04), implying greater mesial-to-lateral spread. Other pairs of localizations were not significantly different (B-H-corrected *p* > 0.05) **(Fig 4B)**. Additionally, an analysis of the effects of different target windows (6s, 11s, 16s, 21s) on HFER localization revealed that the mesial-to-lateral HFER gradient remained consistent for windows up to 16 seconds post-seizure onset, with some diminishing effect observed at 21 seconds **(Supplemental Fig 6).**

**Figure 4.**
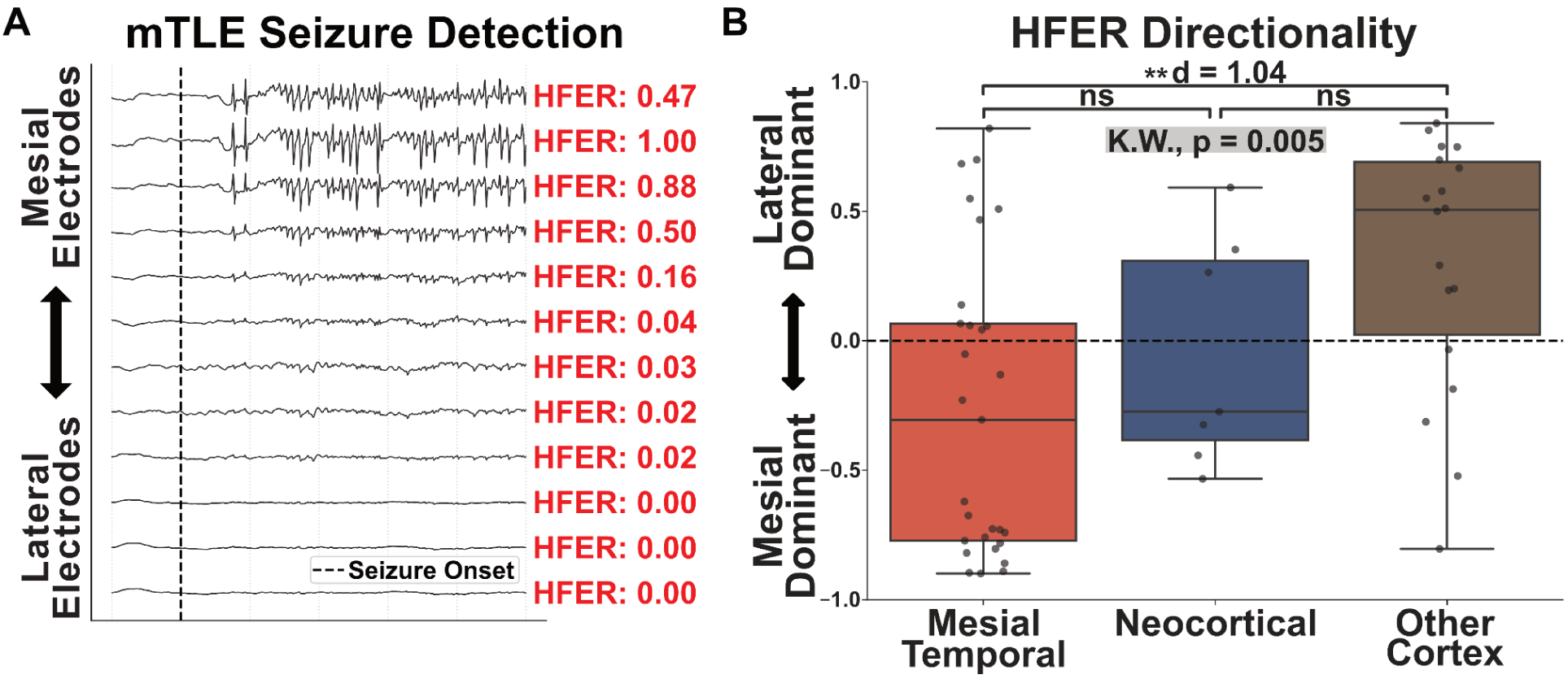
Mesial-to-lateral pattern of seizure HFER across epilepsy localizations. **(A)** Representative iEEG recordings during seizure onset in an mTLE patient. Electrodes are arranged from mesial (top) to lateral (bottom) temporal lobe locations. HFER values for each electrode are shown in red, demonstrating higher values in mesial electrodes. **(B)** Boxplots show the HFER directionality in patients with mTLE, temporal neocortical epilepsy, or other cortical epilepsy. The y-axis shows the Pearson correlation between the morphological feature and the mesial-to-lateral axis. A significant difference (K.W., p = 0.005) is observed. Patients with mTLE have a negative correlation, implying disproportionately more high frequency energy at seizure onset in the mesial structures.. Each dot represents an individual patient, with significant differences between SOZ types indicated by asterisks (* p < 0.05, ** p < 0.01, *** p < 0.001).

### A machine learning algorithm using the mesial-to-lateral pattern of spike and seizure features predicts a clinical diagnosis of mTLE

To understand how well our features distinguish mTLE vs either temporal neocortical or other localizations (combining the latter two localizations into a single class), we trained a set of logistic regression models using a nested LOOCV style architecture. The first model we created was using just the mesial-to-lateral pattern of interictal spike features (spike rate, timing, and morphology). Across all patients from MUSC and HUP, we were able to classify mTLE at a balanced accuracy of 0.71, and an AUC of 0.75 (95% CI: 0.59-0.89) **(Fig 5A)**. Compared to the spike-only model, the seizure-only model, where only the mesial-to-lateral spread of seizure HFER was used to train the model had a balanced accuracy of 0.69 and an AUC of 0.70 (95% CI: 0.55-0.84). The combined model, where both interictal and ictal features were used in training, performed with a balanced accuracy of 0.73 and an AUC of 0.82 (95% CI: 0.70-0.92). The combined model performance was not significantly higher than that of the ictal-only model (two-tailed Delong’s test: *p* = 0.08). In our combined model, the most important feature was spike rate, followed by seizure HFER and morphological features **(Fig 5B)**.

**Figure 5.**
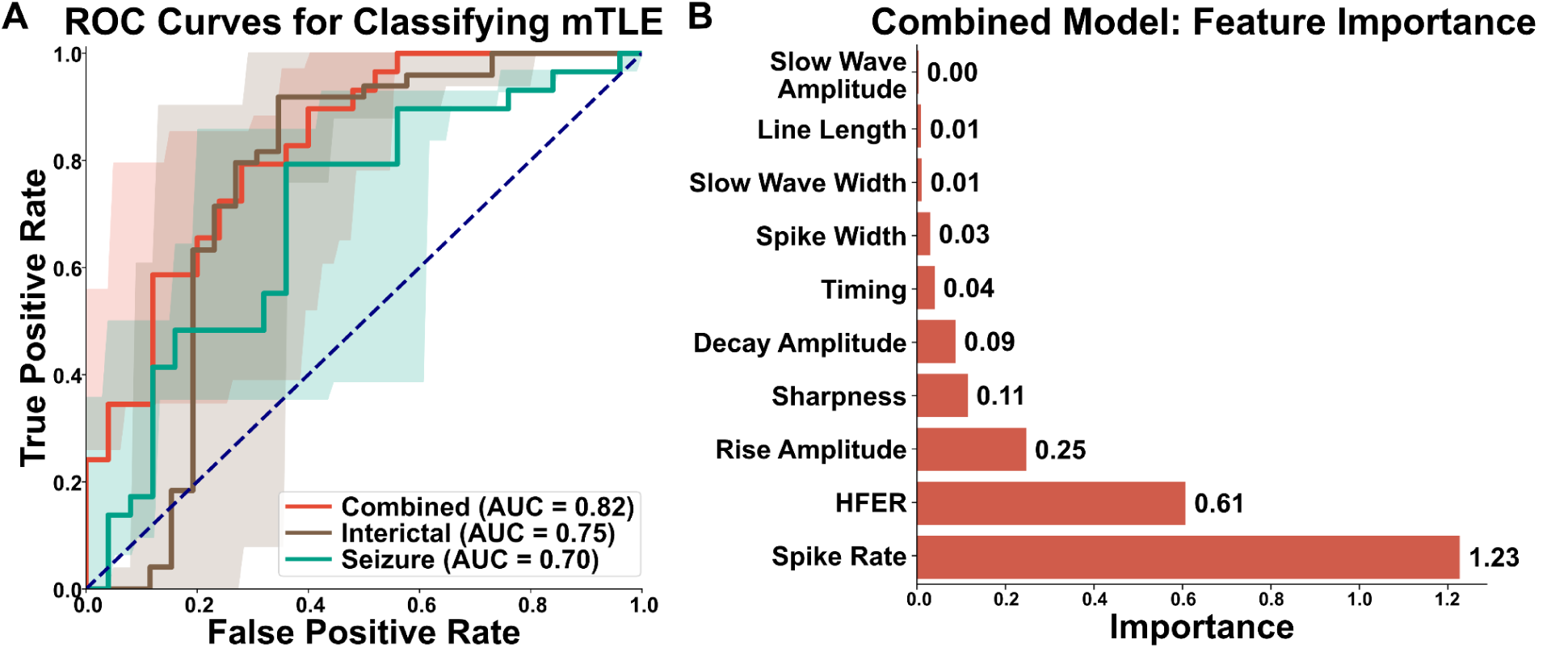
Machine learning classification of mTLE and feature importance analysis. **(A)** Receiver Operating Characteristic (ROC) curves for mTLE classification using different data types. The combined model achieved an AUC of 0.82, compared to AUCs of 0.75 and 0.70 for the interictal-only and seizure-only models respectively, with the difference between the combined and seizure-only models approaching but not reaching statistical significance (Delong’s test, p = 0.08). Shaded areas represent 95% confidence intervals. **(B)** Feature importance in the combined classification model. Bar plot shows the relative importance of various features in distinguishing mTLE. Spike rate emerges as the most influential feature (importance = 1.23), followed by seizure high frequency energy ratio (HFER, importance = 0.61) and rise amplitude (importance = 0.25). Other morphological and timing features contribute to a lesser extent.

### There is no significant difference in the spatial distribution of spike features according to post-mesial temporal ablation 12 month surgical outcome

Of our patient cohort, 20 patients received a laser ablation of the hippocampus or amygdala. We designated good outcomes to be Engel Class I, and anything else as a bad outcome (Jaber et al., 2024). We compared our modeled probability of a localization of mTLE between patients who had a good outcome after undergoing a mesial temporal lobe ablation, those who had a poor outcome, and those who did not undergo a mesial temporal lobe ablation. There was no statistical difference between groups (Kruskall-Wallis *p* = 0.18). There was a trend toward lower median mTLE probabilities for patients who did not receive an mTLE-target ablation (*P* = 0.56 (0.14 - 0.77) than for those who underwent surgery and had either good (*P* = 0.70 (0.58 - 0.88)) or bad outcomes (*P* = 0.67 (0.58 - 0.76)) **(Fig 6A)**. Additionally, we created a separate model to predict good or bad outcomes for patients who underwent mesial temporal lobe ablation using the same LOOCV style architecture. All models performed poorly: the AUC of the interictal model was 0.62 (95% CI: 0.34-0.85), the AUC of the seizure-only model was 0.42 (95% CI: 0.08-0.78), and the AUC of the combined model was 0.51 (95% CI: 0.21-0.80) **(Fig. 6b)**.

**Figure 6.**
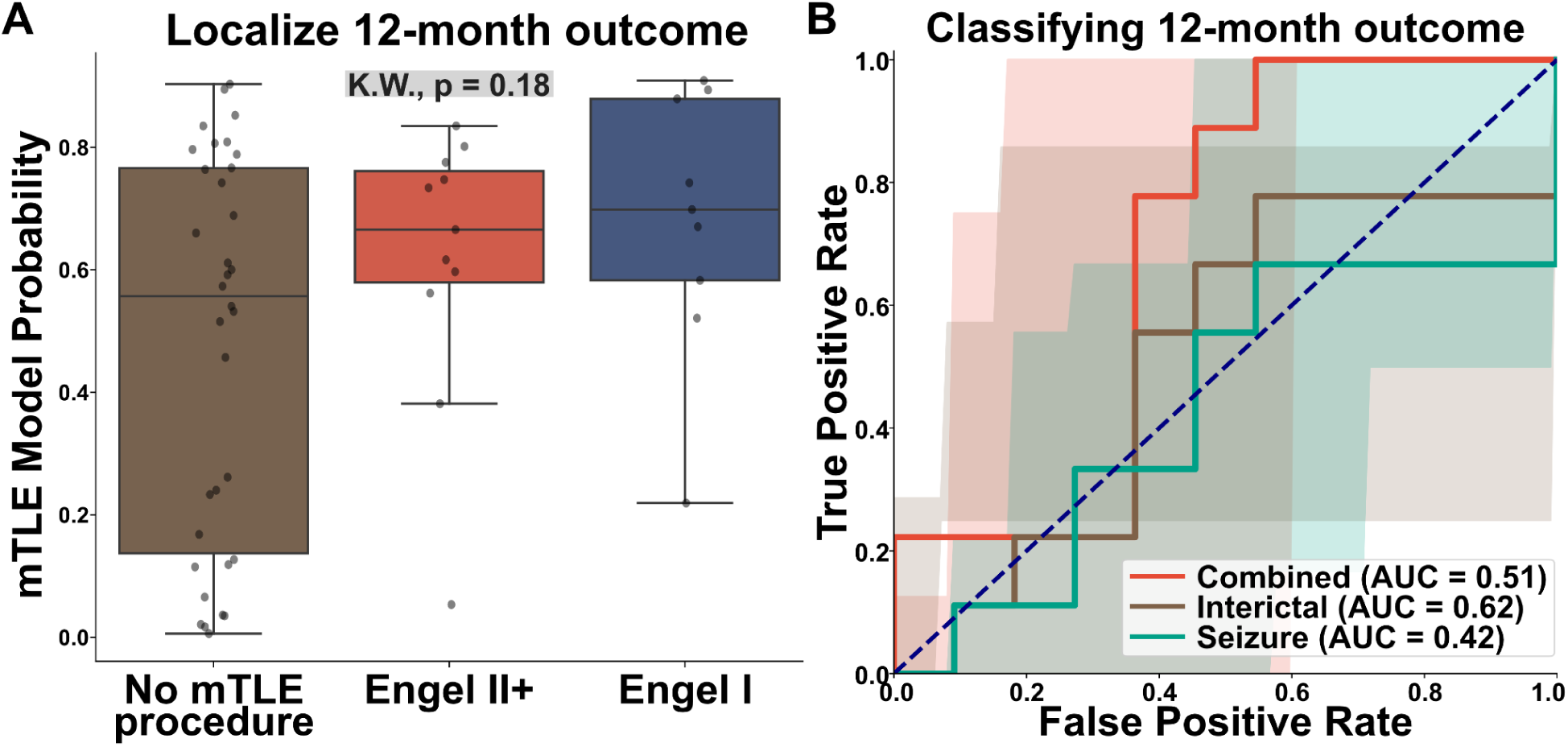
12-month Engel outcome analysis for patients with mesial temporal lobe ablations. **(A)** Box plots showing the distribution of mTLE model probabilities across different surgical outcomes. Patients are grouped by no mTLE procedure, Engel II+ (unfavorable outcome), and Engel I (favorable outcome). No significant difference is observed between groups (Kruskal-Wallis test, p = 0.18). **(B)** ROC curves for classifying 12-month Engel outcomes using different data types. The combined model (AUC = 0.51) performs similarly to chance, while the interictal-only model (AUC = 0.62) shows slightly better performance. The seizure-only model (AUC = 0.42) performs below chance level. Shaded areas represent 95% confidence intervals. This figure suggests that the current mTLE classification model and individual electrophysiological features have limited predictive power for long-term surgical outcomes in mTLE patients.

## Discussion

Here we compared the mesial-to-lateral patterns of spikes and seizures between patients with mTLE and other epilepsy localizations. Leveraging automated detection and analysis of large numbers of spikes, we were able to conduct a detailed examination of spatiotemporal patterns that would have been prohibitively time-consuming with manual methods. We found that the mesial-to-lateral pattern of spike rates and morphology distinguished patients with mTLE from other localizations, in a direction consistent with our hypothesis that spikes in mTLE originate in the mesial temporal lobe and spread laterally. Similar localization was achieved with mesial-to-lateral patterns of seizure energy. A model combining spatiotemporal patterns of spike and seizure features predicted a diagnosis of mTLEs, but did not predict outcome after undergoing ablation of the mesial temporal structures.

### Patients with mTLE have a mesial-to-lateral spread pattern of spike rates and spike morphological features

The regional source of interictal spikes in epilepsy remains a subject of debate. While spike location has been shown to correlate with the epileptogenic cortex (Conrad, Tomlinson, et al., 2020; Dworetzky & Reinsberger, 2011; Kim et al., 2010; Lee et al., 2014; Smith et al., 2022), this relationship is imperfect (Marsh et al., 2010). The concept of spike source localization is based on the assumption that the origin of spikes indicates the epileptogenic zone (Mégevand et al., 2014). However, even when spike location is probed more precisely with intracranial EEG, spikes often appear to arise from multiple sources (Alarcon et al., 1994). For example, in mTLE, spikes are observed in multiple locations, including the lateral neocortex (Merlet & Gotman, 1999; Mitsuhashi et al., 2021; Plummer et al., 2008; Wennberg et al., 2011; Withers et al., 2023). One complicating factor in understanding spike sources is that intracranial EEG recordings are subject to incomplete spatial sampling, potentially capturing only the spread of spikes from unrecorded cortical regions. This limitation could lead to misinterpretation of multifocal spikes: a single unsampled spike origin with diverse propagation patterns might appear as multiple independent sources.

Here, we hypothesized that spikes in mTLE preferentially arise from the mesial temporal lobe structures, leading to a mesial-to-lateral pattern of rate, morphology, and timing that is distinct from other epilepsy localizations. Our hypothesis leads to specific predictions about spikes in mTLE that we tested here: 1) Spike rates should be higher in the mesial temporal electrodes, decreasing moving laterally. 2) Spikes recorded in the mesial temporal electrodes should be higher amplitude and sharper, as they are recording closer to the source of the spike (Hufnagel et al., 2000; Serafini & Loeb, 2015; Thomas et al., 2023). 3) In the case of sequences of spikes occurring in close succession but with discernable latency, the earliest spikes in a sequence should occur towards the mesial temporal lobe. Our results are largely concordant with these predictions, with significantly different mesial-to-lateral patterns of spike rate and morphology between different epilepsy localizations. There was no significant difference in the mesial-to-lateral pattern of spike timing between localizations, but we observed a trend in the expected direction of earlier spikes mesially in patients with mTLE. These results support the hypothesis that spikes preferentially originate in or near the epileptogenic zone in mTLE. Our findings could be consistent with either a single mesial temporal spike source in mTLE or multiple spike sources, with the mesial temporal source predominating. Prior studies in both EEG and magnetoencephalography found evidence supporting multiple distinct spike clusters in epilepsy (Oishi et al., 2006; Tomlinson et al., 2019; Van ’t Ent et al., 2003). A future direction would be to apply a similar technique to cluster spike sequences based on their timing and morphology to understand how often patients with mTLE have multiple independent spike clusters.

### Patients with mTLE have a mesial-to-lateral spread pattern of HFER from seizures

The mesial-to-lateral pattern of seizure HFER differed across epilepsy localizations. Our results suggest that in mTLE patients, mesially located electrode contacts express larger amounts of high frequency energy as compared to lateral electrodes, whereas patients with other localizations demonstrate the reverse, with larger energy values in the temporal electrode contacts. Additionally, our seizure HFER-stability metric (more information in the Supplemental Materials) is largely consistent across seizures within a patient, despite the established variability of seizure spread and severity within patients (Gleichgerrcht et al., 2021; Pattnaik et al., 2023). These findings also support that HFER is a simple quantitative metric to estimate the relative involvement of different electrode contacts in seizure onset.

### A model combining the mesial-to-lateral patterns of spikes and seizures predicts a localization of mTLE

Our model to localize mTLE achieved the highest accuracy when we combined both ictal and interictal features. In our combined model, the spike rate correlation coefficient feature provided the most information on whether a patient had mTLE. Our seizure HFER correlation coefficient for spatial spread provided the second most information for classifying mTLE, and spike morphology features were the next most important after this. The importance of the spike rate feature supports the greater use of spike rate data in addition to seizure information when performing surgical decision making for suspected mTLE.

### The modeled probabilities of mTLE are similar between patients with good and bad outcome after mesial temporal ablations

There was no clear distinction between 12-month good and bad surgical outcomes when using our combined model predicted probability for mTLE. This lack of clear separation between good and bad outcomes may be explained by a combination of factors: First, the low number of patients who underwent ablation (*N* = 20) limited power. Second, outcomes may be influenced by factors beyond the choice of surgical intervention. Mesial temporal lobe ablations often vary in extent and precise location, and incomplete ablations may explain postoperative seizure recurrence even amongst patients with well-localized mTLE (Jermakowicz et al., 2017; Kohlhase et al., 2021; D. D. Liu et al., 2021). Third, missed cases of bilateral mesial temporal lobe epilepsy may explain some cases of postoperative seizure recurrence in mTLE (King-Stephens et al., 2015). These latter two hypotheses for our null result lead to a prediction that some patients with high modeled probability of mTLE would have a poor outcome after a mesial temporal lobe ablation (because they had an incomplete ablation or bilateral foci); conversely, it leads to the prediction that everyone with good outcomes should have high probability of mTLE. Indeed, only one of the nine patients with Engel I outcome after a mesial temporal lobe ablation had a modeled probability of mTLE < 0.5. That patient had a one-year Engel Ib outcome, and then afterward developed recurrent focal impaired awareness seizures approximately two years after surgery. We are unable to draw conclusions from this post-hoc analysis of a single patient combined with the small number of patients with Engel I outcomes, however these results suggest that low modeled probabilities of mTLE may reveal patients unable to benefit from a mesial temporal lobe ablation.

### Limitations

Our study has several limitations. First, the slight variations in mesial-to-lateral temporal electrode placements across patients may introduce inconsistencies in data collection, given that spike morphology is influenced by the geometry and distance from electrode contact to the source of the current (Herreras, 2016). However, this limitation likely increases the probability of Type II errors, suggesting that the observed mesial-to-lateral patterns may be even stronger than our findings indicate. Second, our reliance on a single spike detector may not capture the full spectrum of epileptiform activity. This constraint, similar to the electrode placement issue, potentially reduces the study’s statistical power. Improved spike detectors (Janca et al., 2015; Jing et al., 2020) may yield more accurate detections. Lastly, the limited number of temporal neocortical epilepsy (TNC) patients in our cohort restricts our ability to compare TNC and mTLE with statistical confidence.

### Relevance for clinical translation

Our finding that more frequent, high amplitude, and sharper spikes occur in the mesial temporal structures in mTLE agrees with longstanding clinical intuition (Bautista et al., 1999; Rodin et al., 2009). Our study delivers a method to measure these features *at scale* across the entirety of an intracranial recording, thus supporting the feasibility of using spike rate, morphology, and timing features in surgical planning. To facilitate external validation, we make our code and example spike datasets publicly available. If our results can be validated in large multicenter datasets, they would support integrating quantitative spike analysis in surgical planning, which could 1) improve our ability to localize the epileptogenic cortex, and 2) reduce the number of seizures needed in the EMU, reducing morbidity and cost.

## Conclusion

Our findings indicate that the mesial-to-lateral patterns of the morphology of interictal spikes and seizures vary depending on the epilepsy localization and can predict a localization of mTLE. Univariate analyses suggest that interictal data is just as informative as data from seizures at revealing mTLE localization. This highlights the potential of interictal data, which is often more abundant and easier to collect. Our results suggest that combining both interictal and ictal information could provide a more comprehensive picture. This integrated approach can enhance epileptic diagnosis and assist clinicians in making more informed decisions regarding interventions targeting the mesial temporal lobe.

## Data Availability Statement

Github repository will be released with code to replicate all analysis reported in the study. All data tables will be available for download.

## References

1. Alarcon, G., Guy, C. N., Binnie, C. D., Walker, S. R., Elwes, R. D., & Polkey, C. E. (1994). Intracerebral propagation of interictal activity in partial epilepsy: Implications for source localisation. *Journal of Neurology*, Neurosurgery & Psychiatry, 57(4), 435–449. 10.1136/jnnp.57.4.435

2. Andrews, J. P., Ammanuel, S., Kleen, J., Khambhati, A. N., Knowlton, R., & Chang, E. F. (2020). Early seizure spread and epilepsy surgery: A systematic review. Epilepsia, 61(10), 2163–2172. 10.1111/epi.16668

3. Bartolomei, F., Chauvel, P., & Wendling, F. (2008). Epileptogenicity of brain structures in human temporal lobe epilepsy: A quantified study from intracerebral EEG. Brain, 131(7), 1818–1830. 10.1093/brain/awn111

4. Bautista, R. E. D., Cobbs, M. A., Spencer, D. D., & Spencer, S. S. (1999). Prediction of Surgical Outcome by Interictal Epileptiform Abnormalities During Intracranial EEG Monitoring in Patients with Extrahippocampal Seizures. Epilepsia, 40(7), 880–890. 10.1111/j.1528-1157.1999.tb00794.x

5. Bink, H., Sedigh-Sarvestani, M., Fernandez-Lamo, I., Kini, L., Ung, H., Kuzum, D., Vitale, F., Litt, B., & Contreras, D. (2018). Spatiotemporal evolution of focal epileptiform activity from surface and laminar field recordings in cat neocortex. Journal of Neurophysiology, 119(6), 2068–2081. 10.1152/jn.00764.2017

6. Brown, M. W., Porter, B. E., Dlugos, D. J., Keating, J., Gardner, A. B., Storm, P. B., & Marsh, E. D. (2007). Comparison of novel computer detectors and human performance for spike detection in intracranial EEG. Clinical Neurophysiology, 118(8), 1744–1752. 10.1016/j.clinph.2007.04.017

7. Cai, F., Wang, K., Zhao, T., Wang, H., Zhou, W., & Hong, B. (2022). BrainQuake: An Open-Source Python Toolbox for the Stereoelectroencephalography Spatiotemporal Analysis. Frontiers in Neuroinformatics, 15. 10.3389/fninf.2021.773890

8. Conrad, E. C., Bernabei, J. M., Kini, L. G., Shah, P., Mikhail, F., Kheder, A., Shinohara, R. T., Davis, K. A., Bassett, D. S., & Litt, B. (2020). The sensitivity of network statistics to incomplete electrode sampling on intracranial EEG. Network Neuroscience, 4(2), 484. 10.1162/netn_a_00131

9. Conrad, E. C., Tomlinson, S. B., Wong, J. N., Oechsel, K. F., Shinohara, R. T., Litt, B., Davis, K. A., & Marsh, E. D. (2020). Spatial distribution of interictal spikes fluctuates over time and localizes seizure onset. Brain, 143(2), 554–569. 10.1093/brain/awz386

10. Davis, K. A., Devries, S. P., Krieger, A., Mihaylova, T., Minecan, D., Litt, B., Wagenaar, J. B., & Stacey, W. C. (2018). The effect of increased intracranial EEG sampling rates in clinical practice. Clinical Neurophysiology, 129(2), 360–367. 10.1016/j.clinph.2017.10.039

11. Donos, C., Breier, J., Friedman, E., Rollo, P., Johnson, J., Moss, L., Thompson, S., Thomas, M., Hope, O., Slater, J., & Tandon, N. (2018). Laser ablation for mesial temporal lobe epilepsy: Surgical and cognitive outcomes with and without mesial temporal sclerosis. Epilepsia, 59(7), 1421–1432. 10.1111/epi.14443

12. Dworetzky, B. A., & Reinsberger, C. (2011). The role of the interictal EEG in selecting candidates for resective epilepsy surgery. Epilepsy & Behavior, 20(2), 167–171. 10.1016/j.yebeh.2010.08.025

13. Engel, J. (1993). Update on surgical treatment of the epilepsies. Summary of the Second International Palm Desert Conference on the Surgical Treatment of the Epilepsies (1992). Neurology, 43(8), 1612–1617. 10.1212/wnl.43.8.1612

14. Engel, J., McDermott, M. P., Wiebe, S., Langfitt, J. T., Stern, J. M., Dewar, S., Sperling, M. R., Gardiner, I., Erba, G., Fried, I., Jacobs, M., Vinters, H. V., Mintzer, S., Kieburtz, K., & Early Randomized Surgical Epilepsy Trial (ERSET) Study Group, for the. (2012). Early Surgical Therapy for Drug-Resistant Temporal Lobe Epilepsy: A Randomized Trial. JAMA, 307(9), 922–930. 10.1001/jama.2012.220

15. Epilepsy (ILAE) 1997–2001, C. on N. of the I. L. A., Wieser, H. G., Blume, W. T., Fish, D., Goldensohn, E., Hufnagel, A., King, D., Sperling, M. R., Lüders, H., & Pedley, T. A. (2001). Proposal for a New Classification of Outcome with Respect to Epileptic Seizures Following Epilepsy Surgery. Epilepsia, 42(2), 282–286. 10.1046/j.1528-1157.2001.35100.x

16. Gleichgerrcht, E., Greenblatt, A. S., Kellermann, T. S., Rowland, N., Vandergrift, W. A., Edwards, J., Davis, K. A., & Bonilha, L. (2021). Patterns of seizure spread in temporal lobe epilepsy are associated with distinct white matter tracts. Epilepsy Research, 171, 106571. 10.1016/j.eplepsyres.2021.106571

17. Gompel, J. J. V., Meyer, F. B., Marsh, W. R., Lee, K. H., & Worrell, G. A. (2010). Stereotactic electroencephalography with temporal grid and mesial temporal depth electrode coverage: Does technique of depth electrode placement affect outcome? Journal of Neurosurgery, 113(1), 32. 10.3171/2009.12.JNS091073

18. Gross, R. E., Stern, M. A., Willie, J. T., Fasano, R. E., Saindane, A. M., Soares, B. P., Pedersen, N. P., & Drane, D. L. (2018). Stereotactic Laser Amygdalohippocampotomy for Mesial Temporal Lobe Epilepsy. Annals of Neurology, 83(3), 575–587. 10.1002/ana.25180

19. Hamer, H. M., Morris, H. H., Mascha, E. J., Karafa, M. T., Bingaman, W. E., Bej, M. D., Burgess, R. C., Dinner, D. S., Foldvary, N. R., Hahn, J. F., Kotagal, P., Najm, I., Wyllie, E., & Lüders, H. O. (2002). Complications of invasive video-EEG monitoring with subdural grid electrodes. Neurology, 58(1), 97–103. 10.1212/wnl.58.1.97

20. Hamer, H. M., Najm, I., Mohamed, A., & Wyllie, E. (1999). Interictal epileptiform discharges in temporal lobe epilepsy due to hippocampal sclerosis versus medial temporal lobe tumors. Epilepsia, 40(9), 1261–1268. 10.1111/j.1528-1157.1999.tb00856.x

21. Helmstaedter, C. (2008). Temporal lobe resection—Does the prospect of seizure freedom outweigh the cognitive risks? Nature Clinical Practice Neurology, 4(2), 66–67. 10.1038/ncpneuro0657

22. Herreras, O. (2016). Local Field Potentials: Myths and Misunderstandings. Frontiers in Neural Circuits, 10, 101. 10.3389/fncir.2016.00101

23. Hufnagel, A., Dümpelmann, M., Zentner, J., Schijns, O., & Elger, C. E. (2000). Clinical Relevance of Quantified Intracranial Interictal Spike Activity in Presurgical Evaluation of Epilepsy. Epilepsia, 41(4), 467–478. 10.1111/j.1528-1157.2000.tb00191.x

24. Jaber, K., Avigdor, T., Mansilla, D., Ho, A., Thomas, J., Abdallah, C., Chabardes, S., Hall, J., Minotti, L., Kahane, P., Grova, C., Gotman, J., & Frauscher, B. (2024). A spatial perturbation framework to validate implantation of the epileptogenic zone. Nature Communications, 15(1), 5253. 10.1038/s41467-024-49470-z

25. Janca, R., Jezdik, P., Cmejla, R., Tomasek, M., Worrell, G. A., Stead, M., Wagenaar, J., Jefferys, J. G. R., Krsek, P., Komarek, V., Jiruska, P., & Marusic, P. (2015). Detection of interictal epileptiform discharges using signal envelope distribution modelling: Application to epileptic and non-epileptic intracranial recordings. Brain Topography, 28(1), 172–183. 10.1007/s10548-014-0379-1

26. Jermakowicz, W. J., Kanner, A. M., Sur, S., Bermudez, C., D’Haese, P.-F., Kolcun, J. P. G., Cajigas, I., Li, R., Millan, C., Ribot, R., Serrano, E. A., Velez, N., Lowe, M. R., Rey, G. J., & Jagid, J. R. (2017). Laser thermal ablation for mesiotemporal epilepsy: Analysis of ablation volumes and trajectories. Epilepsia, 58(5), 801–810. 10.1111/epi.13715

27. Jing, J., Sun, H., Kim, J. A., Herlopian, A., Karakis, I., Ng, M., Halford, J. J., Maus, D., Chan, F., Dolatshahi, M., Muniz, C., Chu, C., Sacca, V., Pathmanathan, J., Ge, W., Dauwels, J., Lam, A., Cole, A. J., Cash, S. S., & Westover, M. B. (2020). Development of Expert-Level Automated Detection of Epileptiform Discharges During Electroencephalogram Interpretation. JAMA Neurology, 77(1), 103–108. 10.1001/jamaneurol.2019.3485

28. Johnson, E. L. (2019). Seizures and Epilepsy. The Medical Clinics of North America, 103(2), 309–324. 10.1016/j.mcna.2018.10.002

29. Khoo, H. M., von Ellenrieder, N., Zazubovits, N., He, D., Dubeau, F., & Gotman, J. (2018). The spike onset zone. Neurology, 91(7), e666–e674. 10.1212/WNL.0000000000005998

30. Kim, D. W., Kim, H. K., Lee, S. K., Chu, K., & Chung, C. K. (2010). Extent of neocortical resection and surgical outcome of epilepsy: Intracranial EEG analysis. Epilepsia, 51(6), 1010–1017. 10.1111/j.1528-1167.2010.02567.x

31. King-Stephens, D., Mirro, E., Weber, P. B., Laxer, K. D., Van Ness, P. C., Salanova, V., Spencer, D. C., Heck, C. N., Goldman, A., Jobst, B., Shields, D. C., Bergey, G. K., Eisenschenk, S., Worrell, G. A., Rossi, M. A., Gross, R. E., Cole, A. J., Sperling, M. R., Nair, D. R., … Morrell, M. J. (2015). Lateralization of mesial temporal lobe epilepsy with chronic ambulatory electrocorticography. Epilepsia, 56(6), 959–967. 10.1111/epi.13010

32. Kohlhase, K., Zöllner, J. P., Tandon, N., Strzelczyk, A., & Rosenow, F. (2021). Comparison of minimally invasive and traditional surgical approaches for refractory mesial temporal lobe epilepsy: A systematic review and meta-analysis of outcomes. Epilepsia, 62(4), 831–845. 10.1111/epi.16846

33. Lee, C., Kim, J. S., Jeong, W., & Chung, C. K. (2014). Usefulness of interictal spike source localization in temporal lobe epilepsy: Electrocorticographic study. Epilepsy Research, 108(3), 448–458. 10.1016/j.eplepsyres.2013.12.008

34. Liu, D. D., Lauro, P. M., Phillips III, R. K., Leary, O. P., Zheng, B., Roth, J. L., Blum, A. S., Segar, D. J., & Asaad, W. F. (2021). Two-trajectory laser amygdalohippocampotomy: Anatomic modeling and initial seizure outcomes. Epilepsia, 62(10), 2344–2356. 10.1111/epi.17019

35. Liu, Y.-C., Lin, C.-C. K., Tsai, J.-J., & Sun, Y.-N. (2013). Model-Based Spike Detection of Epileptic EEG Data. *Sensors (Basel*, Switzerland), 13(9), 12536–12547. 10.3390/s130912536

36. Lüders, H. O., Najm, I., Nair, D., Widdess-Walsh, P., & Bingman, W. (2006). The epileptogenic zone: General principles. Epileptic Disorders: International Epilepsy Journal with Videotape, 8 *Suppl 2*, S1–9.

37. Marsh, E. D., Peltzer, B., Brown III, M. W., Wusthoff, C., Storm Jr, P. B., Litt, B., & Porter, B. E. (2010). Interictal EEG spikes identify the region of electrographic seizure onset in some, but not all, pediatric epilepsy patients. Epilepsia, 51(4), 592–601. 10.1111/j.1528-1167.2009.02306.x

38. Mégevand, P., Spinelli, L., Genetti, M., Brodbeck, V., Momjian, S., Schaller, K., Michel, C. M., Vulliemoz, S., & Seeck, M. (2014). Electric source imaging of interictal activity accurately localises the seizure onset zone. *Journal of Neurology*, Neurosurgery & Psychiatry, 85(1), 38–43. 10.1136/jnnp-2013-305515

39. Merlet, I., & Gotman, J. (1999). Reliability of dipole models of epileptic spikes. Clinical Neurophysiology, 110(6), 1013–1028. 10.1016/S1388-2457(98)00062-5

40. Mitsuhashi, T., Sonoda, M., Sakakura, K., Jeong, J., Luat, A. F., Sood, S., & Asano, E. (2021). Dynamic tractography-based localization of spike sources and animation of spike propagations. Epilepsia, 62(10), 2372–2384. 10.1111/epi.17025

41. Oishi, M., Kameyama, S., Masuda, H., Tohyama, J., Kanazawa, O., Sasagawa, M., & Otsubo, H. (2006). Single and Multiple Clusters of Magnetoencephalographic Dipoles in Neocortical Epilepsy: Significance in Characterizing the Epileptogenic Zone. Epilepsia, 47(2), 355–364. 10.1111/j.1528-1167.2006.00428.x

42. Pattnaik, A. R., Ghosn, N. J., Ong, I. Z., Revell, A. Y., Ojemann, W. K. S., Scheid, B. H., Georgostathi, G., Bernabei, J. M., Conrad, E. C., Sinha, S. R., Davis, K. A., Sinha, N., & Litt, B. (2023). The seizure severity score: A quantitative tool for comparing seizures and their response to therapy. Journal of Neural Engineering, 20(4), 046026. 10.1088/1741-2552/aceca1

43. Plummer, C., Harvey, A. S., & Cook, M. (2008). EEG source localization in focal epilepsy: Where are we now? Epilepsia, 49(2), 201–218. 10.1111/j.1528-1167.2007.01381.x

44. Ramantani, G., Westover, M. B., Gliske, S., Sarnthein, J., Sarma, S., Wang, Y., Baud, M. O., Stacey, W. C., & Conrad, E. C. (2023). Passive and active markers of cortical excitability in epilepsy. Epilepsia, 64 Suppl 3(Suppl 3), S25–S36. 10.1111/epi.17578

45. Revell, A. Y., Pattnaik, A. R., Conrad, E., Sinha, N., Scheid, B. H., Lucas, A., Bernabei, J. M., Beckerle, J., Stein, J. M., Das, S. R., Litt, B., & Davis, K. A. (2022). A Taxonomy of Seizure Spread Patterns, Speed of Spread, and Associations With Structural Connectivity (p. 2022.10.24.513577). bioRxiv. 10.1101/2022.10.24.513577

46. Rodin, E., Constantino, T., Rampp, S., & Wong, P. K. (2009). Spikes and Epilepsy. Clinical EEG and Neuroscience, 40(4), 288–299. 10.1177/155005940904000411

47. Seidenberg, M., Hermann, B., Wyler, A. R., Davies, K., Dohan Jr., F. C., & Leveroni, C. (1998). Neuropsychological outcome following anterior temporal lobectomy in patients with and without the syndrome of mesial temporal lobe epilepsy. Neuropsychology, 12(2), 303–316. 10.1037/0894-4105.12.2.303

48. Serafini, R. (2019). Similarities and differences between the interictal epileptiform discharges of green-spikes and red-spikes zones of human neocortex. Clinical Neurophysiology, 130(3), 396–405. 10.1016/j.clinph.2018.12.011

49. Serafini, R., & Loeb, J. A. (2015). Enhanced slow waves at the periphery of human epileptic foci. Clinical Neurophysiology: Official Journal of the International Federation of Clinical Neurophysiology, 126(6), 1117–1123. 10.1016/j.clinph.2014.08.023

50. Smith, E. H., Liou, J., Merricks, E. M., Davis, T., Thomson, K., Greger, B., House, P., Emerson, R. G., Goodman, R., McKhann, G. M., Sheth, S., Schevon, C., & Rolston, J. D. (2022). Human interictal epileptiform discharges are bidirectional traveling waves echoing ictal discharges. eLife, 11, e73541. 10.7554/eLife.73541

51. Spencer, S., & Huh, L. (2008). Outcomes of epilepsy surgery in adults and children. The Lancet Neurology, 7(6), 525–537. 10.1016/S1474-4422(08)70109-1

52. Sperling, M. R. (1997). Clinical challenges in invasive monitoring in epilepsy surgery. Epilepsia, 38 *Suppl 4*, S6–12. 10.1111/j.1528-1157.1997.tb04541.x

53. Thomas, J., Kahane, P., Abdallah, C., Avigdor, T., Zweiphenning, W. J. E. M., Chabardes, S., Jaber, K., Latreille, V., Minotti, L., Hall, J., Dubeau, F., Gotman, J., & Frauscher, B. (2023). A Subpopulation of Spikes Predicts Successful Epilepsy Surgery Outcome. Annals of Neurology, 93(3), 522–535. 10.1002/ana.26548

54. Tomlinson, S. B., Wong, J. N., Conrad, E. C., Kennedy, B. C., & Marsh, E. D. (2019). Reproducibility of interictal spike propagation in children with refractory epilepsy. Epilepsia, 60(5), 898–910. 10.1111/epi.14720

55. Vakharia, V. N., Duncan, J. S., Witt, J.-A., Elger, C. E., Staba, R., & Engel Jr, J. (2018). Getting the best outcomes from epilepsy surgery. Annals of Neurology, 83(4), 676–690. 10.1002/ana.25205

56. Van ’t Ent, D., Manshanden, I., Ossenblok, P., Velis, D. N., de Munck, J. C., Verbunt, J. P. A., & Lopes da Silva, F. H. (2003). Spike cluster analysis in neocortical localization related epilepsy yields clinically significant equivalent source localization results in magnetoencephalogram (MEG). Clinical Neurophysiology, 114(10), 1948–1962. 10.1016/S1388-2457(03)00156-1

57. Virtanen, P., Gommers, R., Oliphant, T. E., Haberland, M., Reddy, T., Cournapeau, D., Burovski, E., Peterson, P., Weckesser, W., Bright, J., van der Walt, S. J., Brett, M., Wilson, J., Millman, K. J., Mayorov, N., Nelson, A. R. J., Jones, E., Kern, R., Larson, E., … van Mulbregt, P. (2020). SciPy 1.0: Fundamental algorithms for scientific computing in Python. Nature Methods, 17(3), 261–272. 10.1038/s41592-019-0686-2

58. Wennberg, R., Valiante, T., & Cheyne, D. (2011). EEG and MEG in mesial temporal lobe epilepsy: Where do the spikes really come from? Clinical Neurophysiology, 122(7), 1295–1313. 10.1016/j.clinph.2010.11.019

59. Wiebe, S., Blume, W. T., Girvin, J. P., & Eliasziw, M. (2001). A Randomized, Controlled Trial of Surgery for Temporal-Lobe Epilepsy. New England Journal of Medicine, 345(5), 311–318. 10.1056/NEJM200108023450501

60. Withers, C. P., Diamond, J. M., Yang, B., Snyder, K., Abdollahi, S., Sarlls, J., Chapeton, J. I., Theodore, W. H., Zaghloul, K. A., & Inati, S. K. (2023). Identifying sources of human interictal discharges with travelling wave and white matter propagation. Brain: A Journal of Neurology, awad 259. 10.1093/brain/awad259

