## Supplementary material for "Mesial-to-lateral patterns of epileptiform activity identify the seizure onset zone in mesial temporal lobe epilepsy"

### Supplemental Methods

**Spike and Morphology Detection**
Spikes were detected using an in-house, previously validated automatic detector (Brown et al., 2007; Conrad et al., 2020). An automated, rule-based morphology annotator was created to identify four feature points of our interictal spikes (Liu et al., 2013; Thomas et al., 2023). The first three feature points highlight the main interictal spike body: spike start, spike peak, and spike end point. The fourth feature point, slow wave end point, highlights the end of the slow-wave portion of the spike. An approximation of spike peak was a direct output of the previously validated detector. Before our morphology detector, all signals were demeaned to center the new mean around zero. We first defined the spike polarity by analyzing the signal values surrounding the approximate spike peak derived from the detector. We searched 10 ms surrounding the detector's spike peak. A positive spike polarity was determined if a majority of the the values surrounding the spike peak were less than the spike peak, and vice versa for determining a negative spike polarity. From here, we corrected the approximate spike peak by setting it to the local minima/maxima (minimum if negative polarity,  maximum if positive polarity) within 10 ms of the approximate spike peak derived from the detector. To determine spike start and spike end point, we searched the surrounding 75 ms of the spike peak for a local minima/maxima (dependent on spike polarity) opposite of the spike peak ("A Glossary of Terms Most Commonly Used by Clinical Electroencephalographers," 1974; Arnal-Real et al., 2021; Kooi, 1966). The slow wave end point was defined as the first time in the EEG signal after the spike end point that returns back to baseline. To determine this, we searched the next 200ms-500ms after spike end point to look for 2 events: (1) the first time the signal value crosses zero (to signify our slow wave has started), and (2) the subsequent time the signal crosses zero (to define the slow wave end point). From these four features points a set of 12 morphological features were derived. Calculations and descriptions of these features are listed in **Table S1** and reference diagrams for our feature point annotations and spike detection are listed in **Fig S1**.

We then performed a limited additional visual validation of 50 random spike morphology detections and 50 random timing detections across all patients, balancing the need to estimate the accuracy of automated feature extraction with the fact that an exhaustive visual validation would be prohibitively time-consuming given the number of features we automatically measured. We followed International Federation of Clinical Neurophysiology criteria as a guide to visually describe spike morphology (Kane et al., 2017; Nascimento & Beniczky, 2023). An accurate detection of the spike peak point was defined as one where a visually large, brief deviation in the iEEG signal is observed. Visual spike start and spike end point detection were defined as the return to baseline signal to the left and right of the spike peak. If a slow wave appeared to be present, slow wave end point detection was visually identified as the end of the after-going iEEG slowing following an interictal spike. Additional visual validation for five random patients is included in the supplement: Six example spike detections and their morphology for five random patients are provided (**Fig S2**). Five spike train detections are also provided for five random patients **(Fig S3)**. The spike, morphology, and spike trains example detections were randomly selected for each patient.

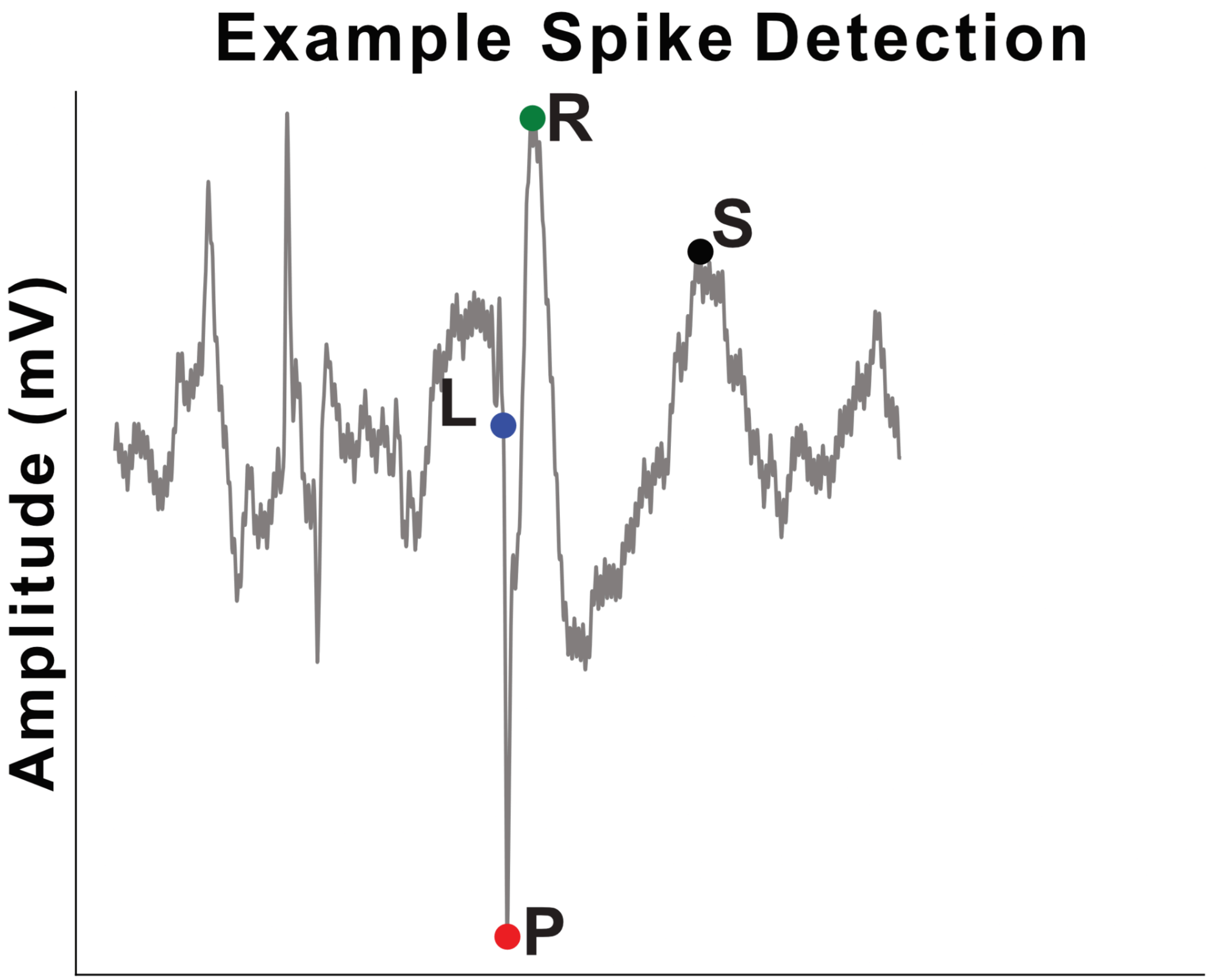


**Figure S1. Example Spike Detection with 4 feature points.** The feature point L is the spike start point, P is the spike peak point, R is the spike end point, and S is the slow wave end point.

| Feature Name | Calculation | Description |
|---|---|---|
| Spike Peak (P) | max(spike wave) | Peak of the sharp portion of spike |

| | | |
|---|---|---|
| Spike Start (L) | min(-75ms from P) | Start of the sharp portion of spike |
| Spike End (R) | min(+75ms from P) | End of the sharp portion of spike |
| Slow Wave End (S) | Second zero crossing (500ms from R) | End of the after going slow-wave following spike |
| Rising Amplitude | P - L | Amplitude of beginning half of spike |
| Decay Amplitude | R - P | Amplitude of returning half of spike |
| Average Amplitude | Rise Amp. + Decay Amp. / 2 | Average of amplitudes of both halves of spike |
| Spike Width | Time between L and R | Duration of spike |
| Rising Width | Time between L and P | Duration of beginning half of spike |
| Decay Width | Time between P and R | Duration of returning half of spike |
| Rise Slope | \| (P - L) / Time between L and P \| | Slope of beginning half of spike |
| Decay Slope | \| (P - L) / Time between L and P \| | Slope of returning half of spike |

**Table S1. Table includes a description and a formula for deriving morphological features.**

**Correlation analysis for choosing morphological features in univariate analysis.** We restricted the number of morphological features we studied in our univariate analyses and machine learning model to prevent Type I errors resulting from multiple testing, and to prevent overfitting. To select features, we visually analyzed the Pearson correlation between features (**Fig. S4**). We made the decision to only look at main spike features (not slow wave features) due to the slow wave features having less accurate detections upon visual validation. To choose which features to keep for univariate analysis, we averaged the pairwise correlations across the rows. Spike width (average $r$ = 0.26), spike sharpness (average $r$ = 0.78), and rising amplitude (average $r$ = 0.74) were the least correlated features from the main spike feature set; these were included in the manuscript morphology univariate analysis. Decay amplitude (average $r$ = 0.75) and line length (average $r$ = 0.76) were not chosen since they had greater average Pearson correlation values. Additionally, hierarchical clustering was performed to understand how each feature captures different aspects of spike morphology. From the main spike features: spike width is in its own group, while spike sharpness and spike rising amplitude branch off early, suggesting these features capture the most distinct information.

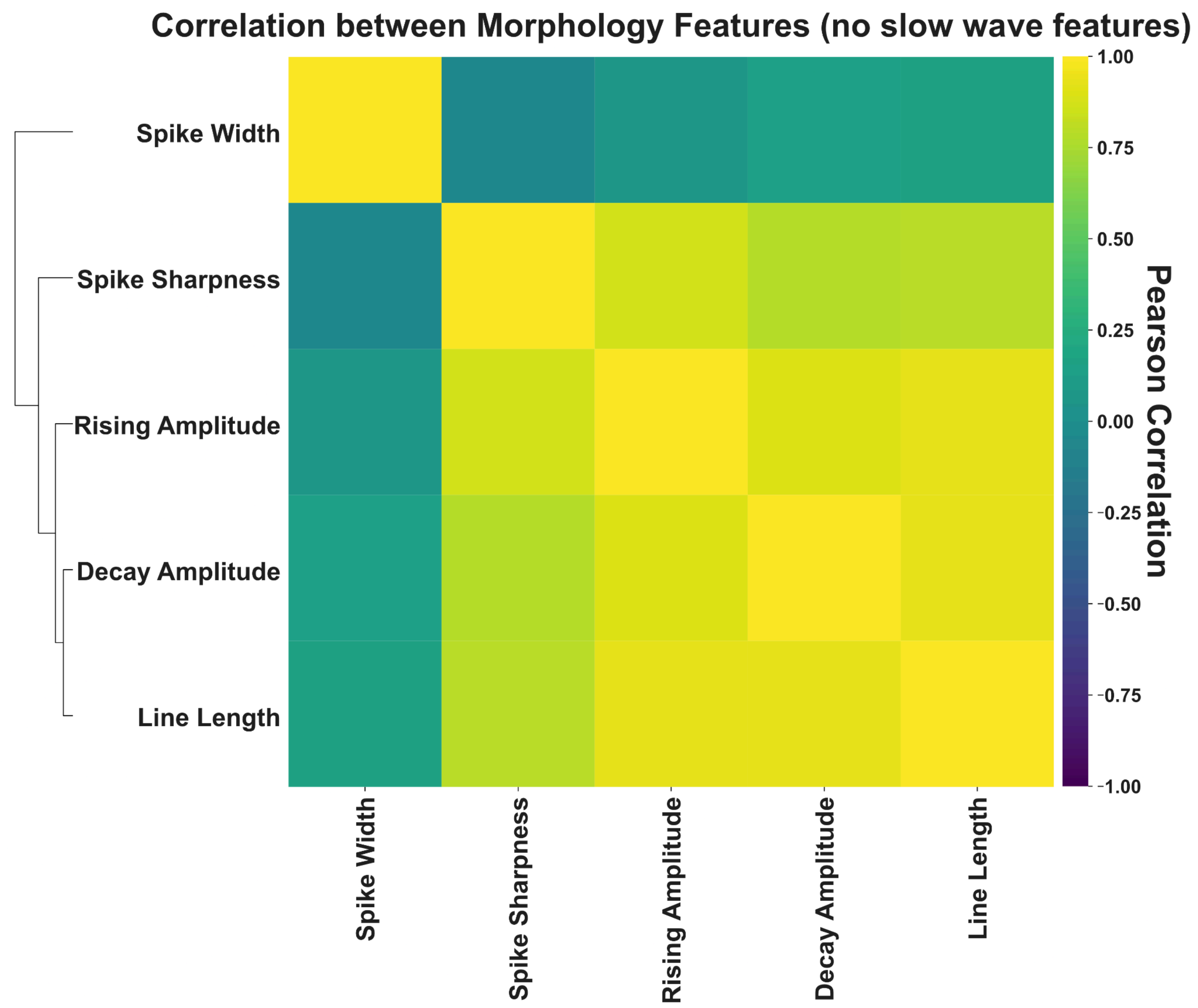


**Figure S4. Correlation between morphological features.** The heatmap displays Pearson correlation coefficients between the main spike morphological features of spikes. Color intensity and scale indicate the strength and direction of correlations. The dendrogram on the left suggests which morphological features branch off first, indicating different aspects of morphology. This analysis provides insights into the relationships between different aspects of spike morphology and may inform feature selection.

**Sequence Stability**

To examine the stability and consistency of spike sequence order over time, we measured the degree preference (DP) across each patient's intracranial recording (Overall DP) and in 30-minute time bins (Segment DP) (Tomlinson et al., 2019). DP measures the preference of an electrode channel's position in a spike sequence, where DP > +20 is labeled "Upstream", DP < -20 is "Downstream", and anywhere in between is "Intermediate". DP was calculated for each contact as the average position of a contact across all spike trains, as calculated in Eq (1), in the specified time bin.

$$(1) \; - \; DP = 100 * (No.\ of\ Downstream - No.\ of\ Upstream) \,/\, (No.\ of\ Total\ Spikes)$$

For each train, the electrode contact containing the earliest occurring spike will have a maximal DP of +100, while the electrode contact containing the last spike in a train will have a minimal DP of -100. To calculate how stable DP is for each patient, the spearman correlation coefficient was calculated between the electrodes in the Segment DP and the Overall DP. To reduce spurious spike activations, only electrode contacts that made up 90% of a patient's total spikes were included in the analysis. To account for sparse time segments, we omitted any segments where less than 25% of the electrodes overlapped with the electrodes included in the Overall DP.

A nonparametric permutation test was used to understand whether our observed DP-stability was statistically significant. To get this new random distribution, DP measures were shuffled randomly across the cohort. This procedure was repeated 1000 times.

**Seizure High Frequency Energy Ratio (HFER)**

To calculate the HFER per channel across seizures and patients, we first identified the seizure onset times. These were determined in a clinical case conference. We next measured the HFER surrounding each seizure onset. We defined a window from 1 second prior to the seizure onset time to 10 seconds after the seizure onset time. This window was chosen to capture the onset and early spread of most seizures. To test the sensitivity of our result to this choice, we also examined additional end times of 5 seconds, 15 seconds, and 20 seconds. We applied a bandpass filter (70-140Hz) to obtain the high frequency component of the signal, and a notch filter (60Hz) to remove line noise and its harmonics. Across the high frequency target window, we calculated energy per channel, defined as the sum of the squared amplitude values of the filtered signal over time. HFER was computed as the ratio of high-frequency (70-140Hz) energy in the target window to that in a baseline window (100-40s pre-seizure). HFER values were normalized using the mean and standard deviation across all channels for each seizure.This was to control for the fact that some seizures had high HFER across all electrodes. We then took the median normalized HFER across seizures, resulting in a single HFER for each electrode.

To determine the effect of our varying target window sizes, we calculated HFER using 5s, 10s, 15s, and 20s end times for our target window sizes. From here we calculated the mesial-to-lateral HFER gradient for all patients, and computed pairwise Pearson correlations between our 4 target window groups.

##### Seizure Stability

To understand the variance between seizures across our cohort, we implemented a method similar to the one used for sequence stability. HFER stability measures the overall likelihood that the electrodes involved in all of a patient's seizures are similarly involved across their individual seizures. To understand how similar seizures were per patient and across the cohort, we calculated the spearman correlation between the overall HFER per electrode across seizures and HFER per electrode in individual seizures for all patients. Overall HFER was defined as the median HFER value per electrode channel across all seizures for a single patient. Seizure HFER was the basic HFER value computed per electrode contact per seizure.

A nonparametric permutation test was used to understand whether our observed HFER-stability was statistically significant. To get this new random distribution, HFER measures were shuffled randomly across the cohort. This procedure was repeated 1000 times.

### Results

##### Spike detector and morphology detector perform well

To ensure that our algorithm for morphological feature point detection was accurate we performed a visual validation (CA). Across our cohort the main spike feature points (spike start [L], spike end [R], and spike peak point [P]) were determined with an accuracy of 94%. Slow wave end point detection (S) was determined with an accuracy of 88%. These

##### Other morphological features localize mTLE

We compared the mesial-to-lateral spread of spike morphology between localizations, predicting higher-amplitude and sharper spikes in the mesial contacts for mTLE, leading to *negative* Pearson correlations across our amplitude-based features. Due to the exploratory nature of this analysis, correction for multiple comparisons were not performed for this analysis. Mann-Whitney U tests revealed that patients with mTLE had significantly different Pearson correlations than patients with other-cortex localizations for decay amplitude (mTLE: r = -0.62 (-0.77 - -0.21) vs. other-cortex: r = -0.24 (-0.47 - 0.15); U = 254, p = 0.008), rising slope (mTLE: r = -0.61 (-0.81 - -0.26) vs. other-cortex: r = -0.09 (-0.42 - 0.23); U = 217, p = 0.002), decay slope (mTLE: r = -0.70 (-0.80 - -0.54) vs. other-cortex: r = -0.10 (-0.58 - 0.26); U = 195, p = 0.0005), and average amplitude (mTLE: r = -0.61 (-0.79 - -0.25) vs. other-cortex: r = -0.27 (-0.49 - 0.19); U = 253, p =

0.008), implying a stronger mesial-to-lateral gradient of spike morphology in mTLE patients, consistent with our hypothesis of preferential spike generation in mesial temporal structures in mTLE. Additionally, rising slope (mTLE: r = -0.61 (-0.81 - -0.26) vs.temporal neocortical: r = -0.09 (-0.57 - 0.11); U = 109, p =0.05) and rising spike width (mTLE: r = 0.11 (-0.31 - 0.57) vs. temporal neocortical: r = -0.46 (-0.71 - 0.22); U = 286, p =0.04) demonstrate significant different Pearson correlations between mTLE and temporal neocortical patients, further indicating a strong mesial-to-lateral gradient of spike morphology in mTLE patients **(Fig S5)**.

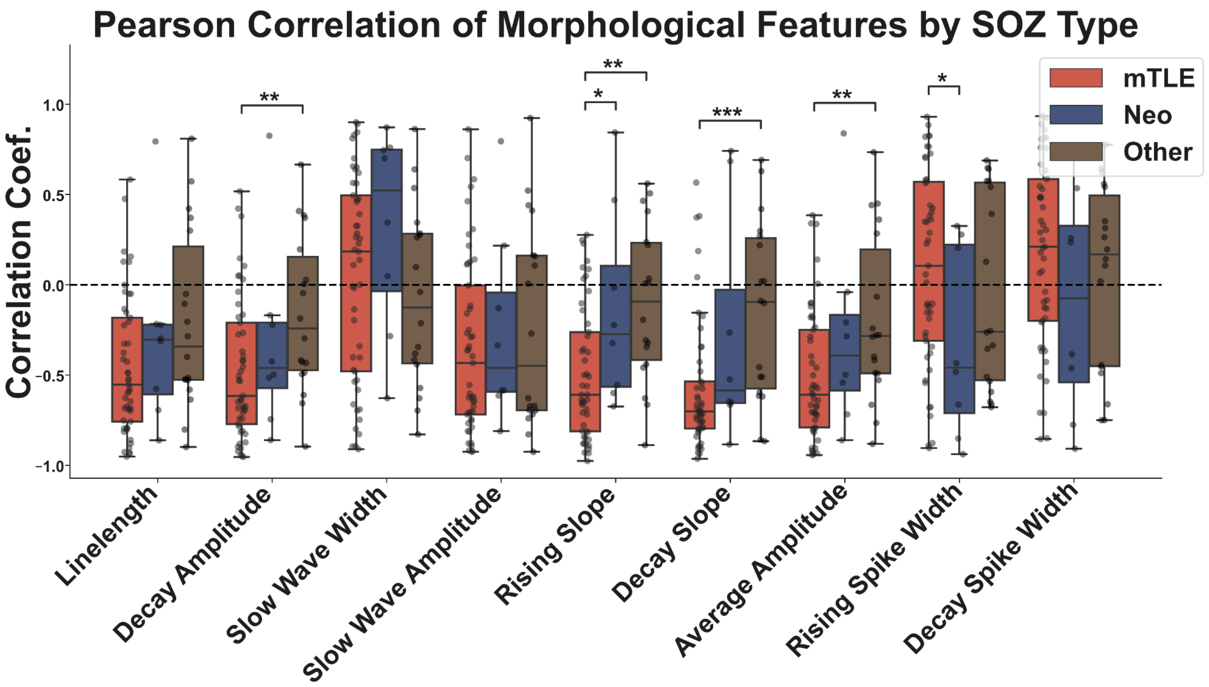


**Figure S5. Pearson correlation coefficients of morphological features across different seizure onset zone (SOZ) types.** Box plots display the distribution of correlation coefficients for nine spike morphology features in mesial temporal lobe epilepsy (mTLE), neocortical (Neo), and other cortical (Other) SOZ types. Each dot represents an individual patient, with significant differences between SOZ types indicated by asterisks (* $p < 0.05$, ** $p < 0.01$, *** $p < 0.001$).

**HFER is robust across varying target window end times.**

Across a representative mTLE seizure, we observe a consistent mesial-to-lateral spread of HFER across all 4 target window sizes **(Fig S6A).** We observe similar spread across all time windows, although the mesial-to-lateral spread started to diminish at the 21 s window. This is further observed in our pairwise Pearson correlation test, where the 21s target window is least correlated with our 6s target window (*r* = 0.68) **(Fig S6B)**. These results suggest stability in the HFER mesial-to-lateral pattern for the first 16 seconds, which then diminishes after this point. Additionally, we tested how well each target window size affects the ability for HFER to localize different epilepsies. Across all window sizes we found a difference in mesial-to-lateral spread between epilepsy localizations (6s: Kruskal-Wallis: $\chi^2(2) = 6.49$, $p = 0.04$; 11s: $\chi^2(2) = 10.49$, $p < 0.01$; 16s: $\chi^2(2) = 8.88$, $p = 0.01$; 21s: $\chi^2(2) = 7.00$, $p = 0.03$) **(Fig S6C).** Post-hoc Mann-Whitney U tests revealed that patients with mTLE had significantly lower Pearson correlations than patients with other-cortex localizations across all target window sizes, implying we are able to capture HFER mesial-to-lateral spread regardless of target window size.

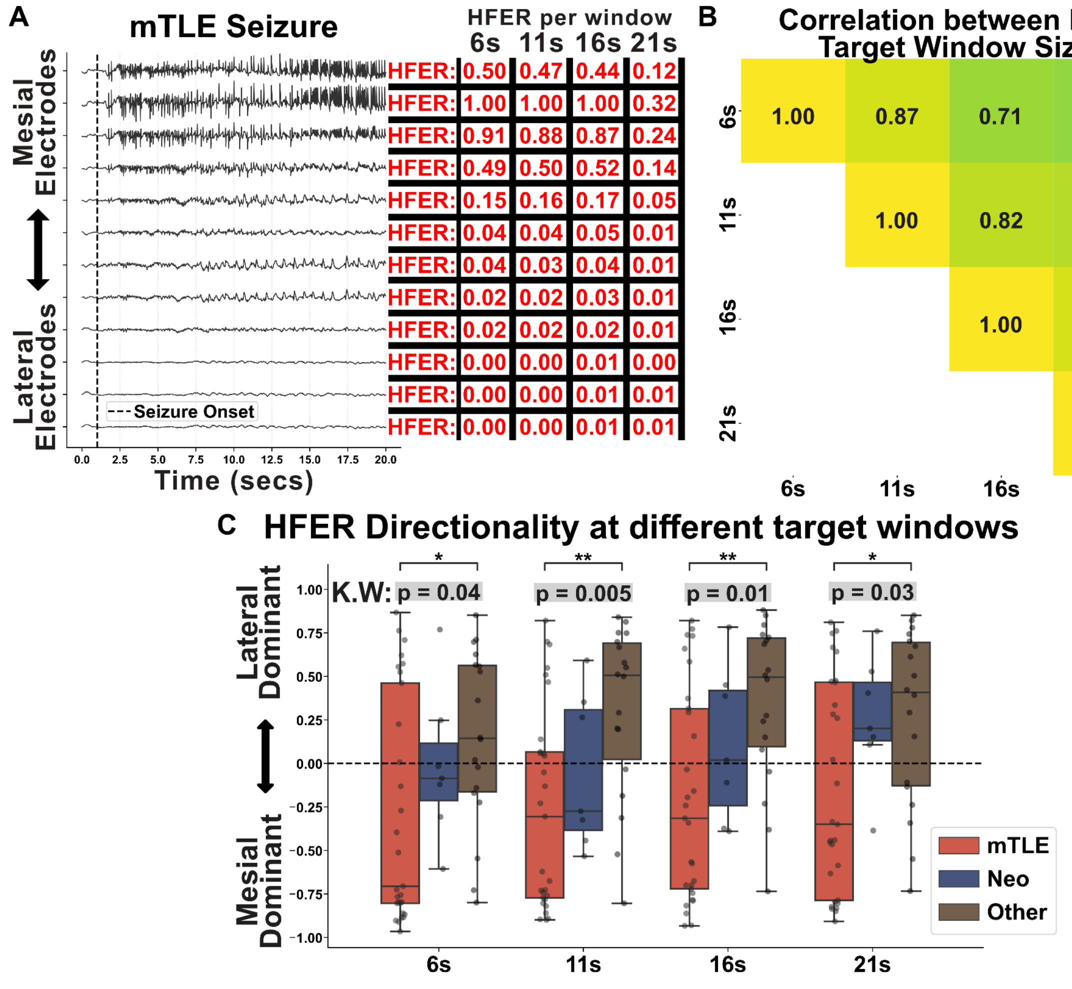


**Figure S6. Analysis of HFER across different time windows. (A)** Representative mTLE seizure recording showing intracranial EEG traces from mesial to lateral electrodes. HFER values are calculated for four different target window sizes (6s, 11s, 16s, 21s) after seizure onset, demonstrating a clear mesial-to-lateral gradient. **(B)** Correlation matrix of HFER values across different target window sizes for the example seizure shown in A. Strong positive correlations are observed between adjacent window sizes (e.g., 6s vs. 11s: 0.87, 11s vs. 16s: 0.82), with decreasing correlation as the difference in window size increases. This analysis suggests that HFER measurements are relatively stable across different time windows, with the 21s window showing the most divergence from shorter windows. **(C)** Box plots display the distribution of correlation coefficients for 4 HFER calculations using different target windows in mesial temporal lobe epilepsy (mTLE), neocortical (Neo), and other cortical (Other) SOZ types, now shown for all patients. Each dot represents an individual patient, with significant differences between SOZ types indicated by asterisks (* $p < 0.05$, ** $p < 0.01$, *** $p < 0.001$). These results suggest that the difference in the mesial-to-lateral pattern of seizure HFER across epilepsy localizations is robust to the choice of time window chosen.

**Spike sequences and seizures are consistent throughout the cohort.**

We also asked whether the ordering of electrode contacts within spike sequences was stable across spike sequences within the same patient. Across all patients, the median [interquartile range (IQR)] of DP stability, a measure of consistency of spike sequence ordering, was 0.51 [0.44 - 0.61] (Nonparametric permutation test, $p = 0.001$), suggesting that spike sequences are largely stable in their propagation order across time. We also measured whether the relative involvement of electrode contacts within seizures as measured by seizure HFER was stable across different seizures. Across all patients, the median [IQR] of seizure HFER stability was 0.61 [0.37 - 0.79] (Nonparametric permutation test, $p = 0.012$), again suggesting stable electrode involvement across seizures.

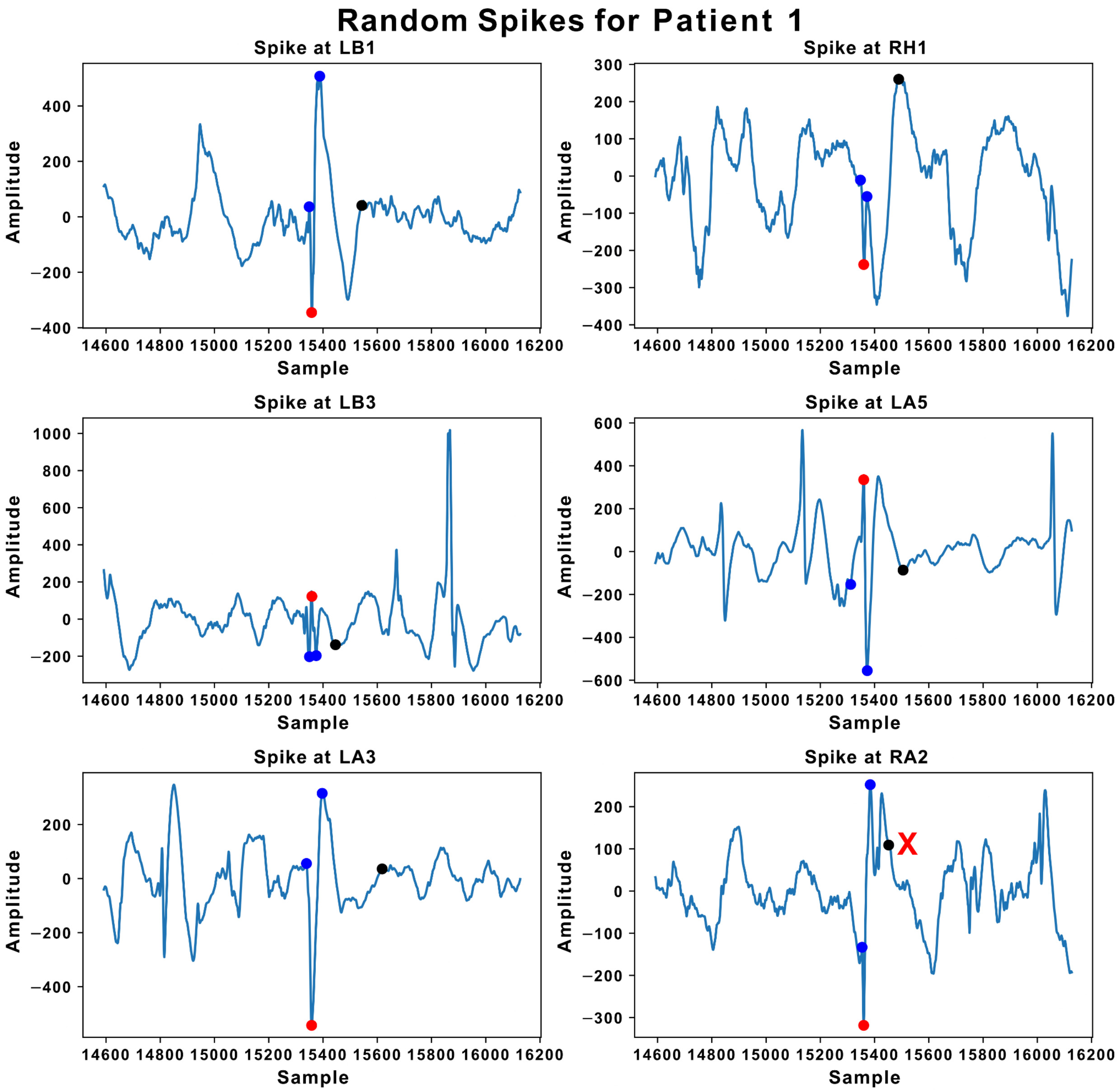

Random Spikes for Patient 1
Spike at LB1
Spike at RH1
Spike at LB3
Spike at LA5
Spike at LA3
Spike at RA2
Amplitude
Sample
X

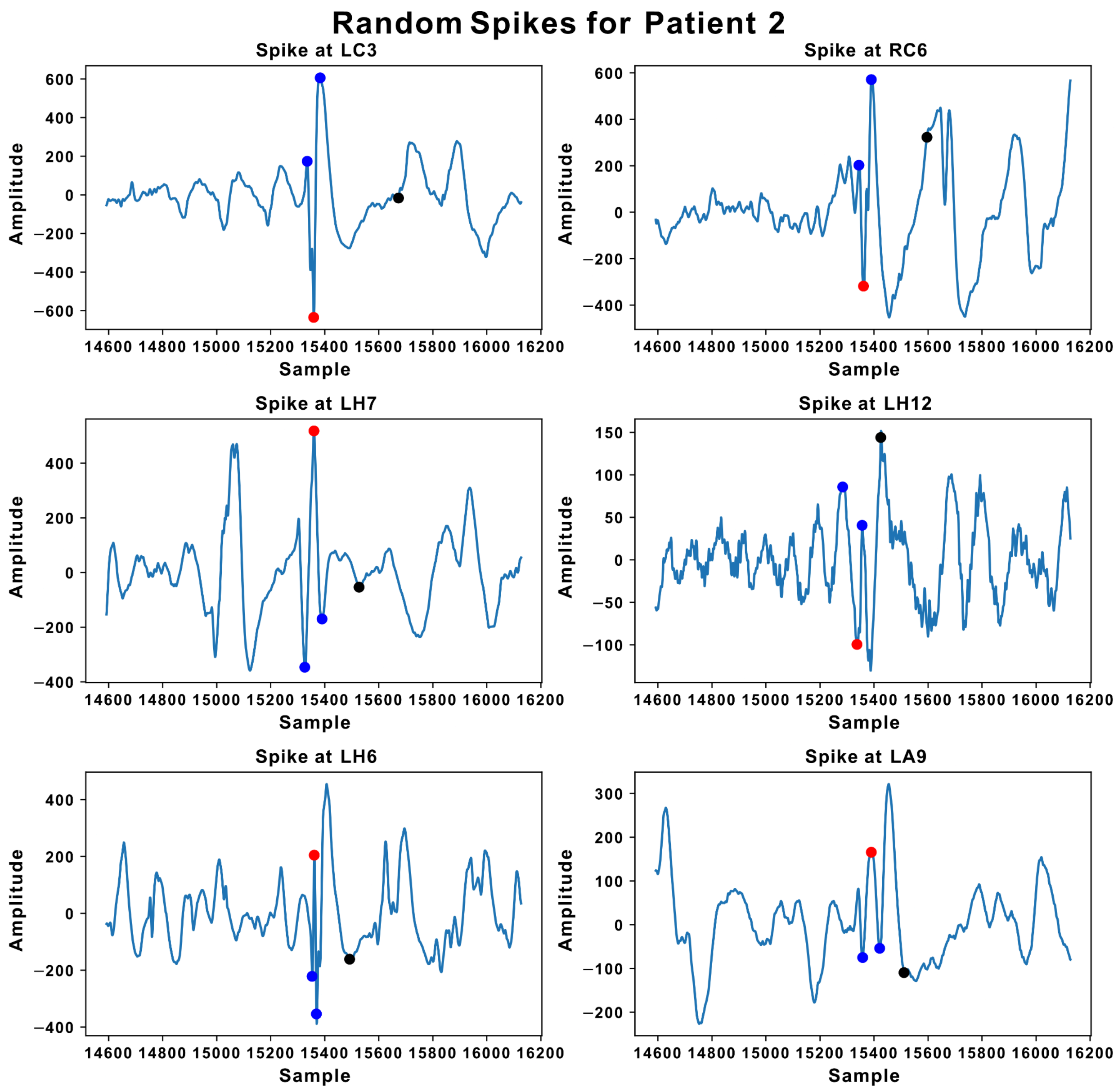

Random Spikes for Patient 2
Spike at LC3
Spike at RC6
Spike at LH7
Spike at LH12
Spike at LH6
Spike at LA9
Amplitude
Sample
14600 14800 15000 15200 15400 15600 15800 16000 16200

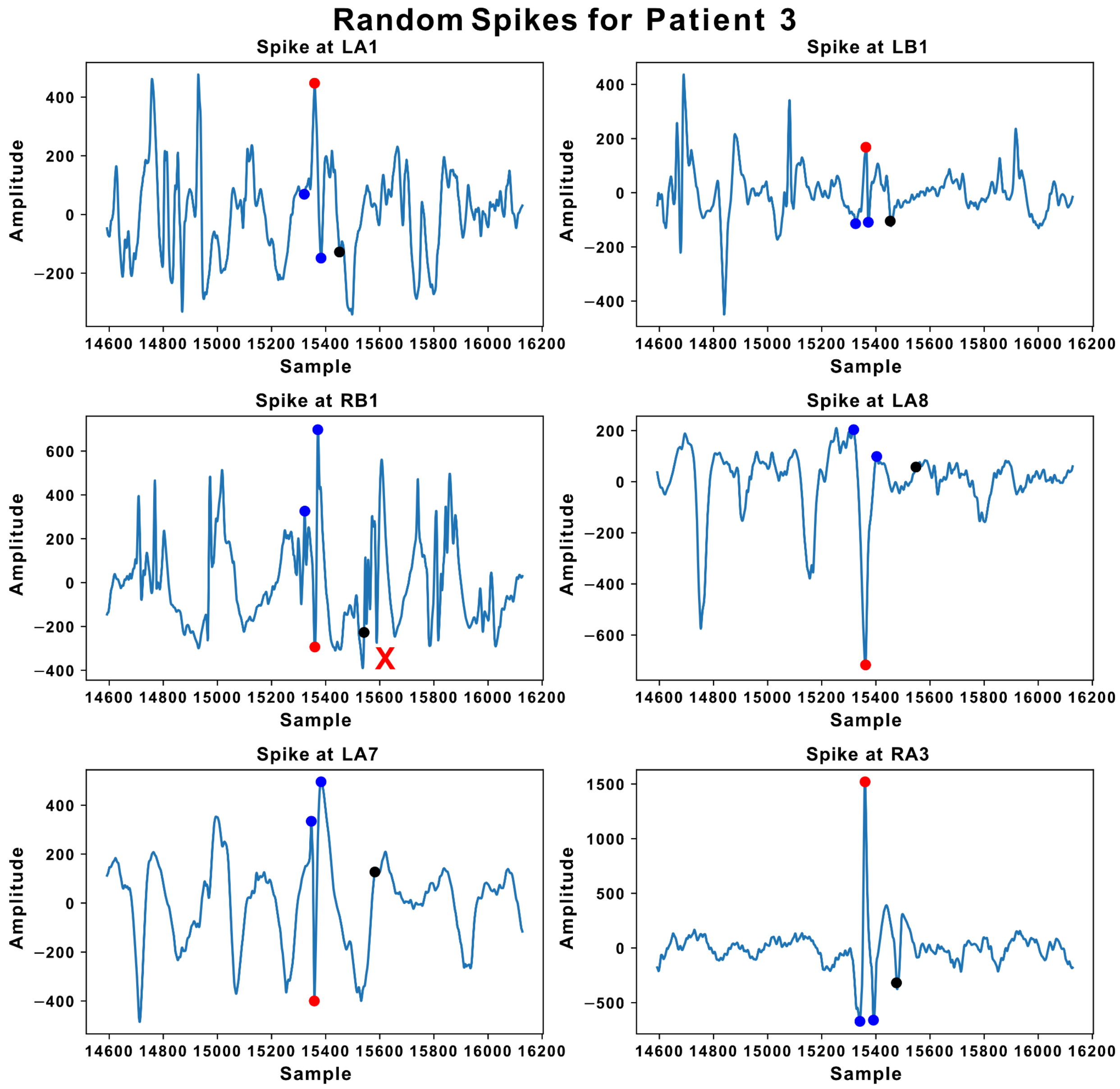

Random Spikes for Patient 3
Spike at LA1
Spike at LB1
Spike at RB1
Spike at LA8
Spike at LA7
Spike at RA3
Amplitude
Sample
X

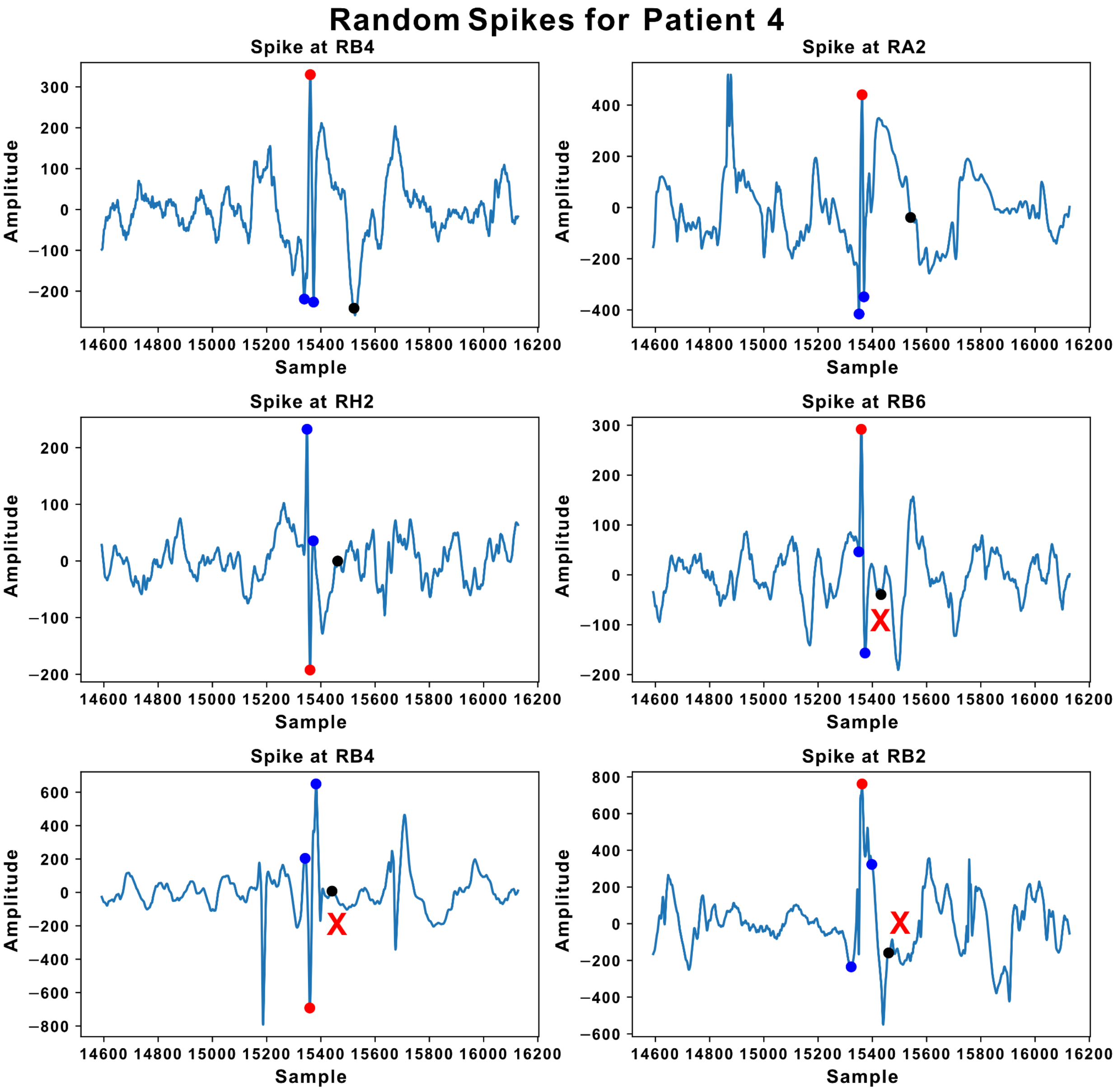

Random Spikes for Patient 4
Spike at RB4
Spike at RA2
Spike at RH2
Spike at RB6
Spike at RB4
Spike at RB2
Amplitude
Sample
X
X
X

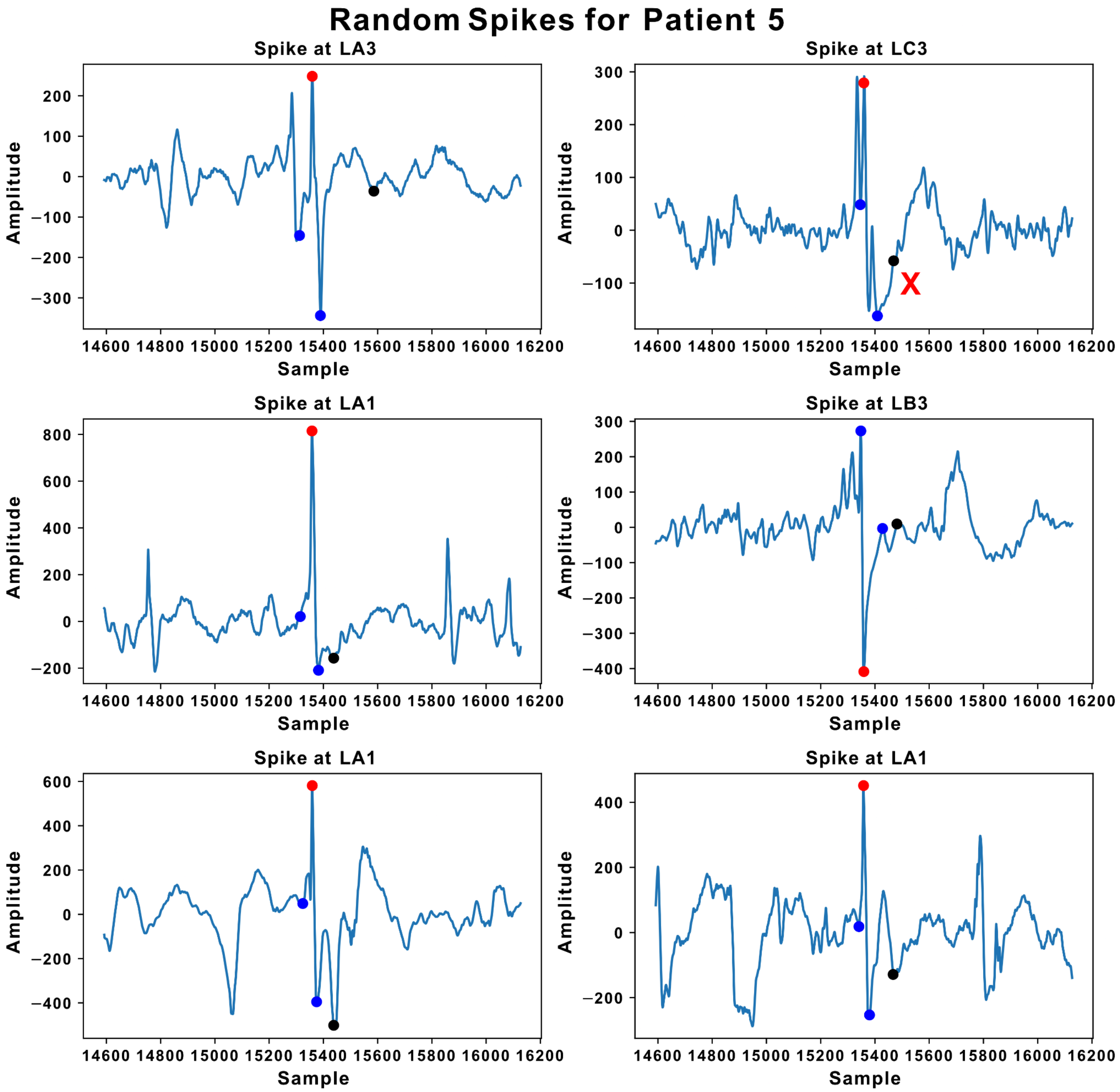


**Figure 2S. Random spike detections + morphology designation for 5 randomly chosen patients.** Each panel contains 6 spike detections and their corresponding morphology according to our automated detector. Each spike highlights a blue dot indicating spike start and end point, a red dot indicating spike peak time, and a black dot indicating the end of the slow wave. From these 4 points, we can perform morphological feature extraction. A red “X” marks annotations that we deemed incorrect.

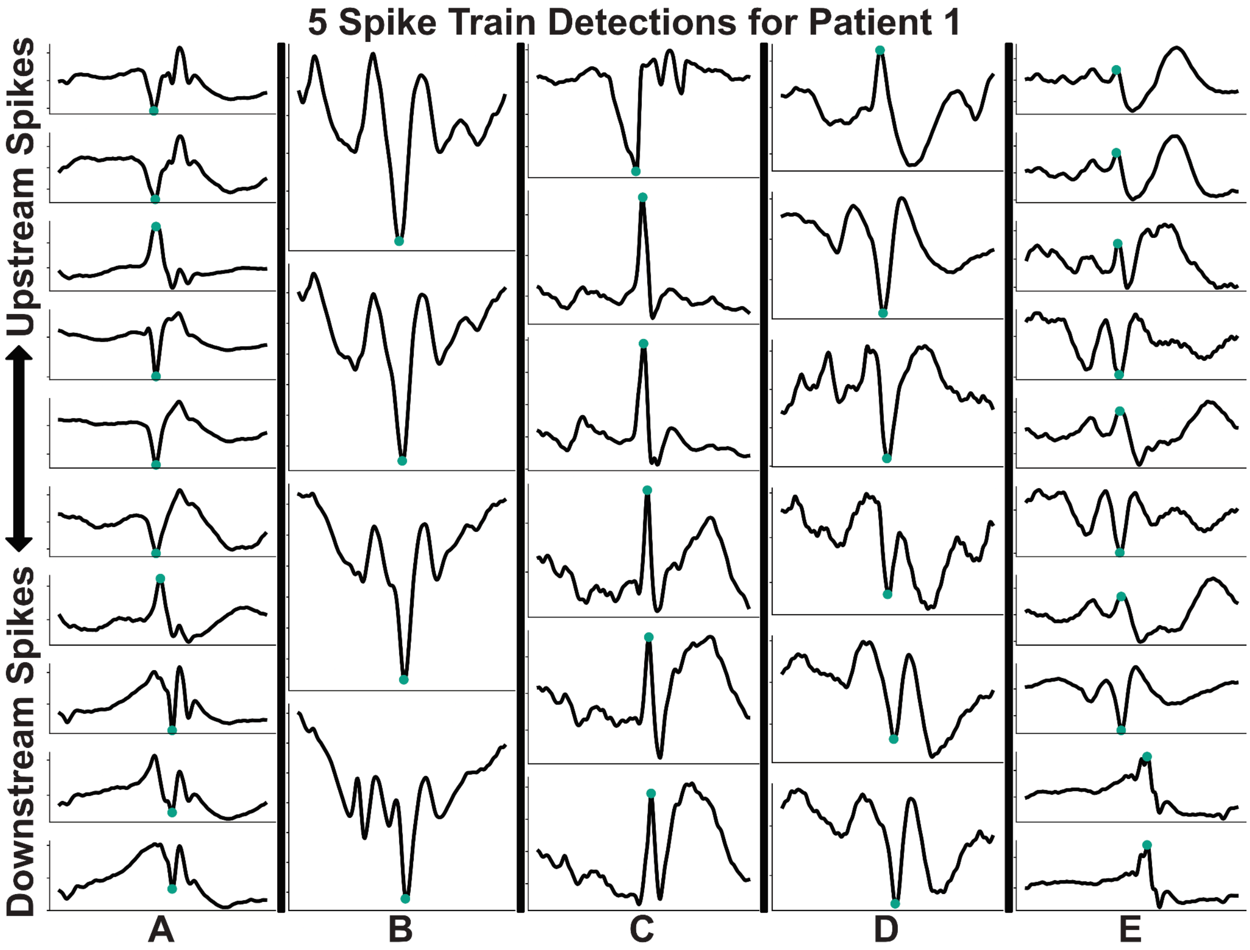

5 Spike Train Detections for Patient 1
Upstream Spikes
Downstream Spikes
A
B
C
D
E

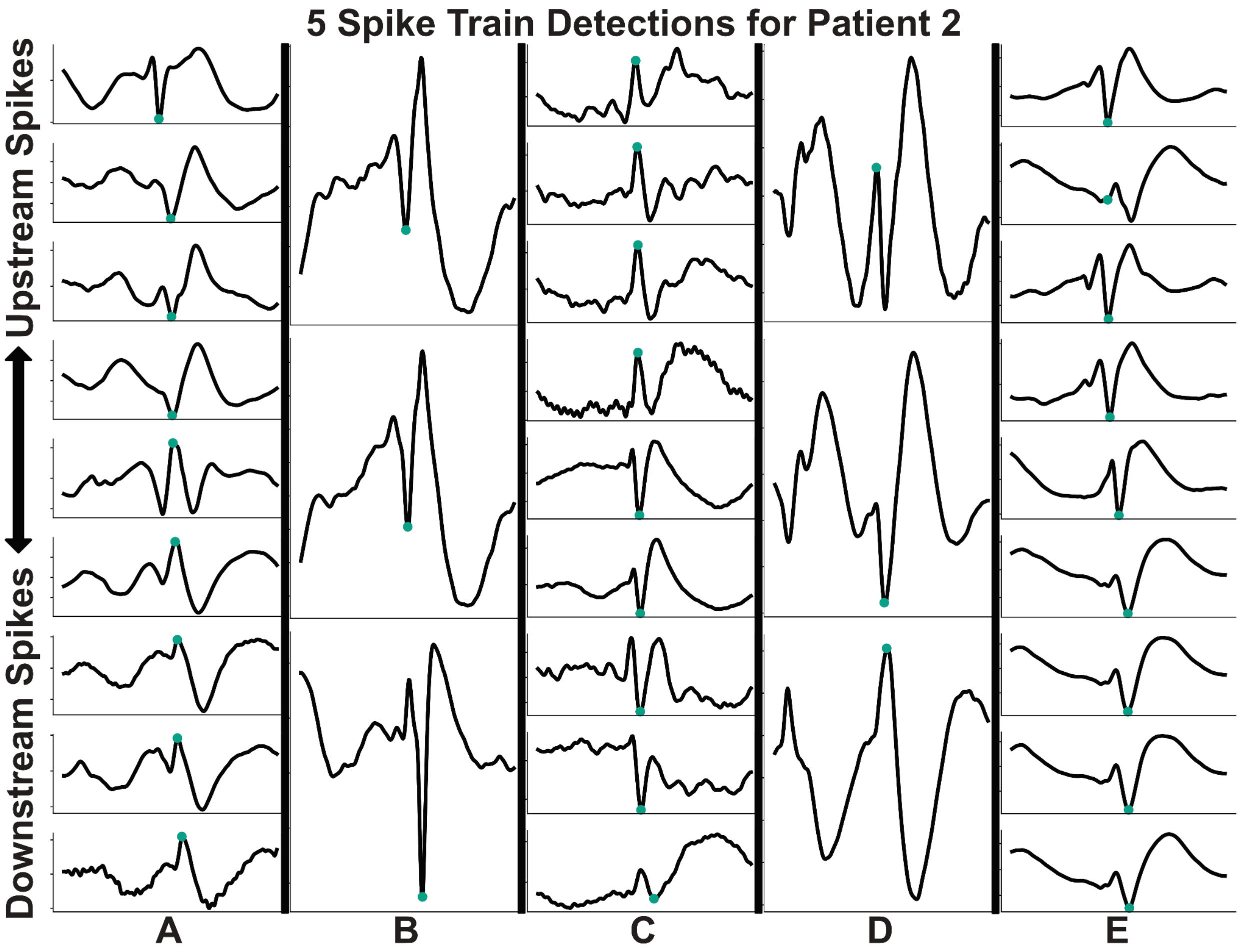

5 Spike Train Detections for Patient 2
Upstream Spikes
Downstream Spikes
A
B
C
D
E

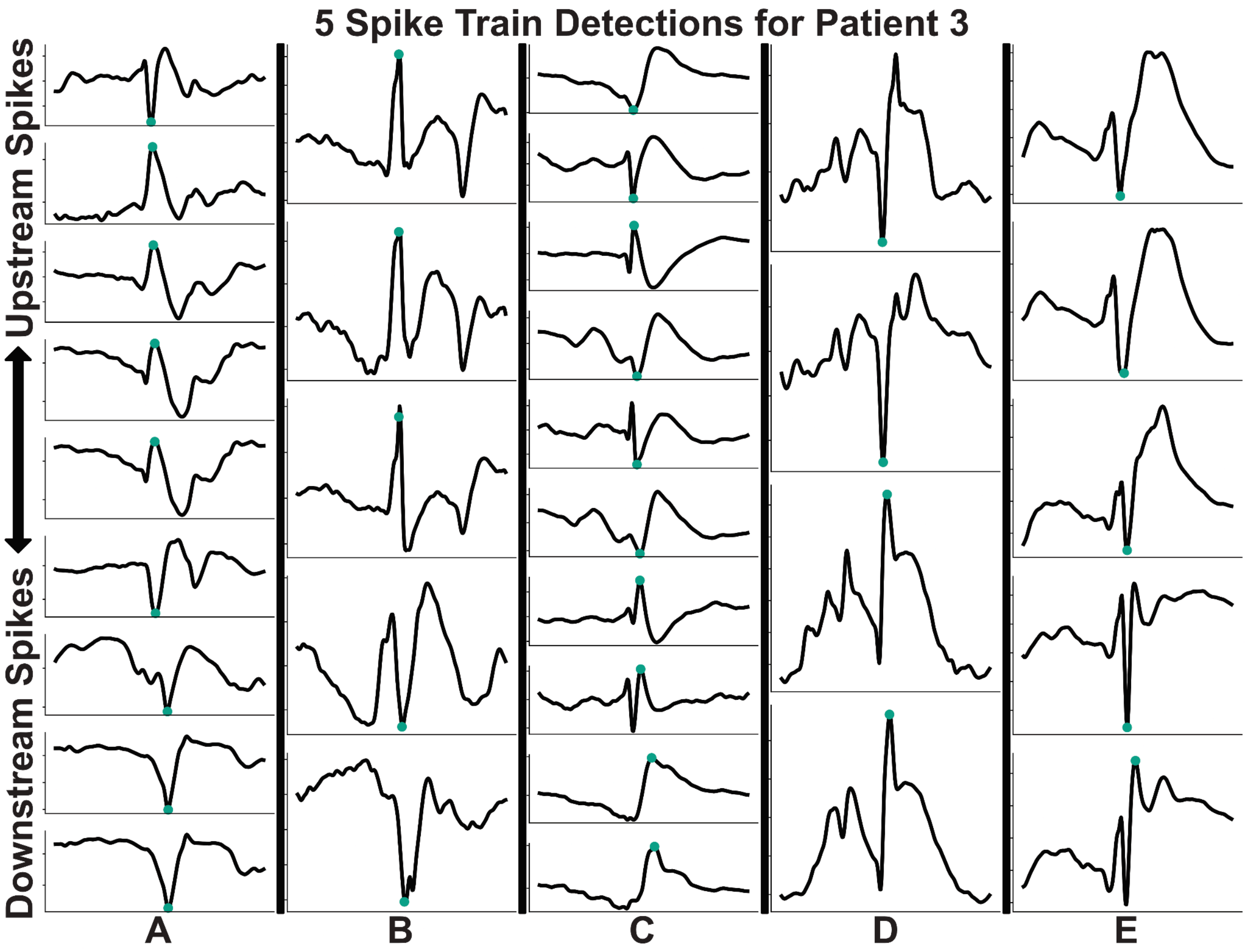

5 Spike Train Detections for Patient 3
Upstream Spikes
Downstream Spikes
A
B
C
D
E

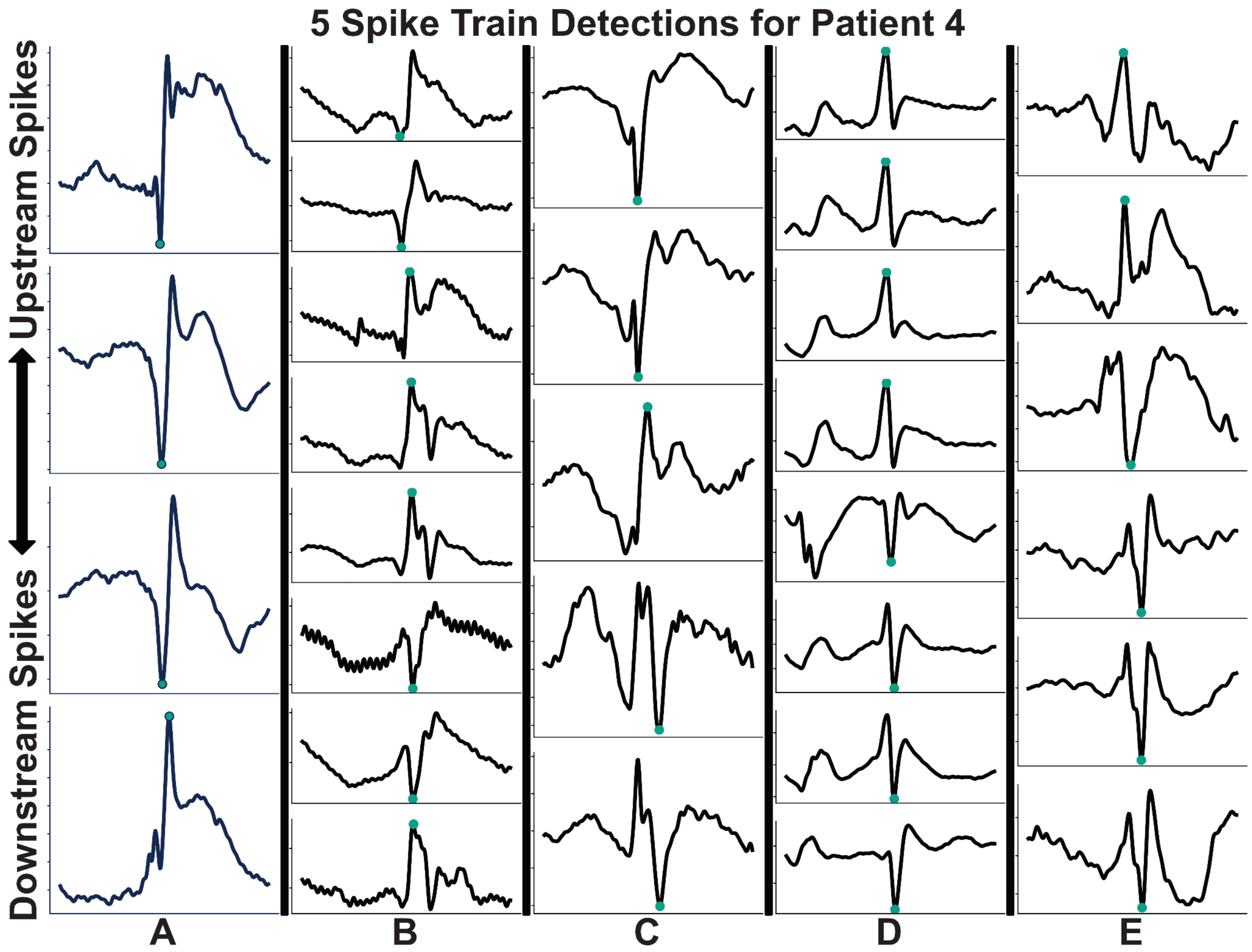
5 Spike Train Detections for Patient 4
Upstream Spikes
Downstream Spikes
A
B
C
D
E

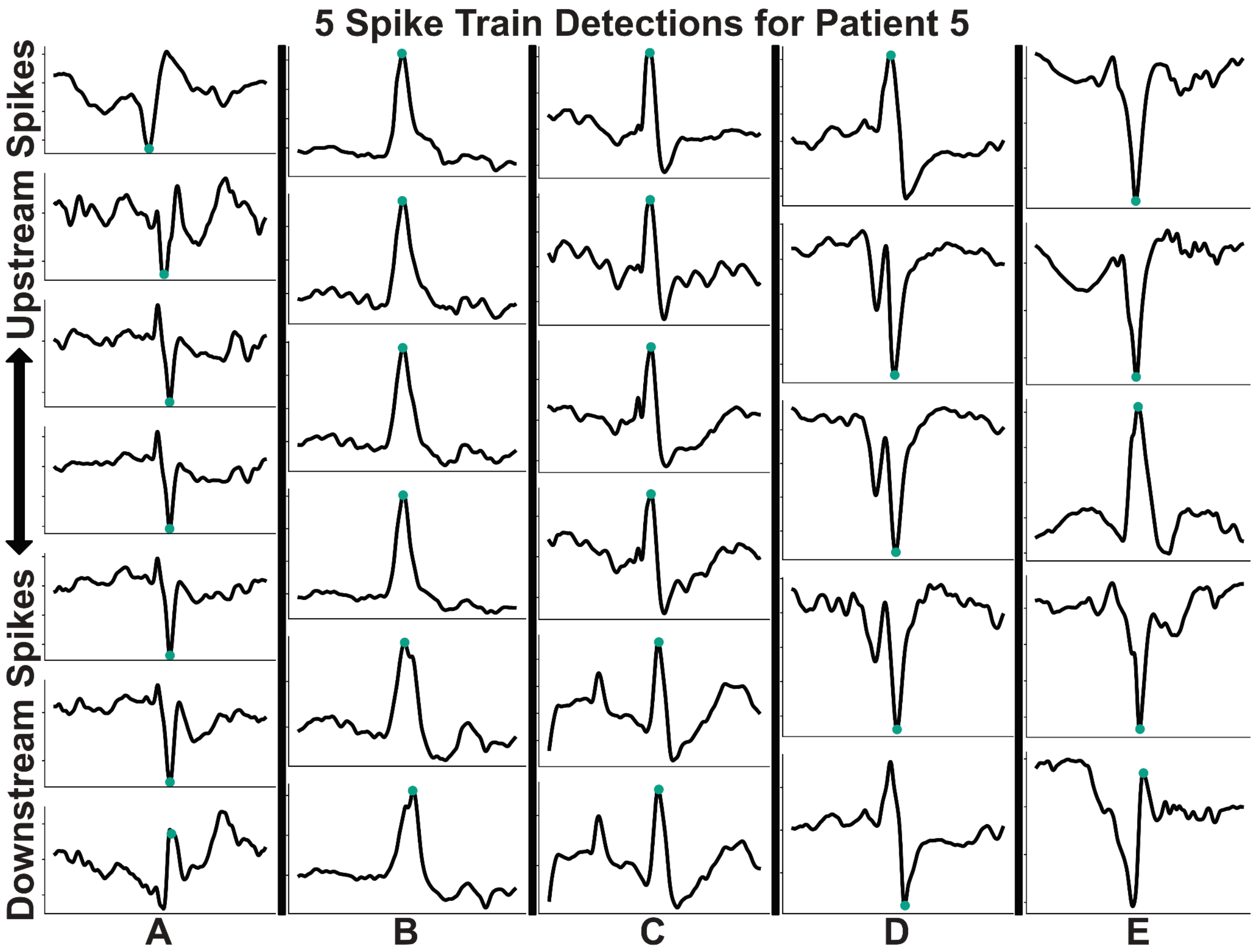


**Figure S3. 5 random spike train detections for 5 randomly chosen patients.** Each panel has 5 random spike train detections labeled (A-E). Spikes trains are displayed vertically, where each spike waveform is a separate spike detected on a new electrode channel in the same train.

#### References


A glossary of terms most commonly used by clinical electroencephalographers: G. E. Chatrian (Chairman), L. Bergamini, M. Dondey, D. W. Klass, M. Lennox-Buchthal and I. Petersén. (1974). *Electroencephalography and Clinical Neurophysiology*, *37*(5), 538–548. https://doi.org/10.1016/0013-4694(74)90099-6

Arnal-Real, C., Mahmoudzadeh, M., Manoochehri, M., Nourhashemi, M., & Wallois, F. (2021). What Triggers the Interictal Epileptic Spike? A Multimodal Multiscale Analysis of the Dynamic of Synaptic and Non-synaptic Neuronal and Vascular Compartments Using Electrical and Optical Measurements. *Frontiers in Neurology*, *12*, 596926. https://doi.org/10.3389/fneur.2021.596926

Brown, M. W., Porter, B. E., Dlugos, D. J., Keating, J., Gardner, A. B., Storm, P. B., & Marsh, E. D. (2007). Comparison of novel computer detectors and human performance for spike detection in intracranial EEG. *Clinical Neurophysiology*, *118*(8), 1744–1752. https://doi.org/10.1016/j.clinph.2007.04.017

Conrad, E. C., Tomlinson, S. B., Wong, J. N., Oechsel, K. F., Shinohara, R. T., Litt, B., Davis, K. A., & Marsh, E. D. (2020). Spatial distribution of interictal spikes fluctuates over time and localizes seizure onset. *Brain*, *143*(2), 554–569. https://doi.org/10.1093/brain/awz386

Kane, N., Acharya, J., Benickzy, S., Caboclo, L., Finnigan, S., Kaplan, P. W., Shibasaki, H., Pressler, R., & van Putten, M. J. A. M. (2017). A revised glossary of terms most commonly used by clinical electroencephalographers and updated proposal for the report format of the EEG findings. Revision 2017. *Clinical Neurophysiology Practice*, *2*, 170–185. https://doi.org/10.1016/j.cnp.2017.07.002

Kooi, K. A. (1966). Voltage‐time characteristics of spikes and other rapid electroencephalographic transients. *Neurology*, *16*(1), 59–59. https://doi.org/10.1212/WNL.16.1.59

Liu, Y.-C., Lin, C.-C. K., Tsai, J.-J., & Sun, Y.-N. (2013). Model-Based Spike Detection of Epileptic EEG Data. *Sensors (Basel, Switzerland)*, *13*(9), 12536–12547. https://doi.org/10.3390/s130912536

Nascimento, F. A., & Beniczky, S. (2023). Teaching the 6 Criteria of the International Federation of Clinical Neurophysiology for Defining Interictal Epileptiform Discharges on EEG Using a Visual Graphic. *Neurology Education*, *2*(2), e200073. https://doi.org/10.1212/NE9.0000000000200073

Thomas, J., Kahane, P., Abdallah, C., Avigdor, T., Zweiphenning, W. J. E. M., Chabardes, S., Jaber, K., Latreille, V., Minotti, L., Hall, J., Dubeau, F., Gotman, J., & Frauscher, B. (2023). A Subpopulation of Spikes Predicts Successful Epilepsy Surgery Outcome. *Annals of Neurology*, *93*(3), 522–535. https://doi.org/10.1002/ana.26548

Tomlinson, S. B., Wong, J. N., Conrad, E. C., Kennedy, B. C., & Marsh, E. D. (2019). Reproducibility of interictal spike propagation in children with refractory epilepsy. *Epilepsia*, *60*(5), 898–910. https://doi.org/10.1111/epi.14720
